# AI in ECG-Based Electrolyte Imbalance Prediction: A Systematic Review and Meta Analysis

**DOI:** 10.1101/2025.01.09.25320300

**Authors:** Muhammed Furkan Dasdelen, Furkan Almas, Zehra Betul Dasdelen, Ayse Nur Balci Yapalak, Carsten Marr

## Abstract

**Background:** Electrolyte imbalances significantly affect heart function, making electrocardiography (ECG) a crucial non-invasive tool. This study systematically reviewed and meta-analyzed AI models’ diagnostic accuracy for detecting these imbalances from ECG, aiming to enhance early detection and improve cardiac care.

**Methods:** We searched 9 databases and reference lists. Two reviewers assessed bias via the Quality Assessment of Diagnostic Accuracy Studies-2 (QUADAS-2). Test performance data were extracted into 2×2 tables and pooled estimates of specificity, sensitivity, and diagnostic odds ratio (DOR) were calculated using a bivariate random-effects model, presented in forest plots and summary receiver operating characteristic curves. We explored heterogeneity by meta-regression, examining internal/external datasets and the number of leads.

**Results:** 21 studies addressing potassium, calcium, and sodium were included. A meta-analysis was conducted only on potassium imbalances (10 studies), covering over 600,000 ECGs from five countries, mostly 12-lead. Among eight studies focused on hyperkalemia, pooled sensitivity, specificity, and DOR were 0.856 (95% CI: 0.829–0.879), 0.788 (0.744–0.826), and 21.8 (17.8–26.7). For hypokalemia (six studies), pooled sensitivity, specificity, and DOR were 0.824 (0.785– 0.856), 0.724 (0.668–0.774), and 12.27 (9.15–16.47). QUADAS-2 assessment showed a 52% high risk of bias in patient selection, mainly due to inadequate sampling details and case-control approaches.

**Conclusion:** AI models can detect ECG-based electrolyte abnormalities, particularly hyperkalemia, and valuable in ICU settings requiring frequent electrolyte assessments and in home monitoring for patients with end-stage renal disease. However, larger retrospective and prospective studies across diverse clinical settings, hospitals, ethnic groups, countries, and regions are warranted.

## Introduction

Electrocardiography (ECG) is a fundamental diagnostic tool used in clinical practice to assess the electrical activity of the heart. It provides crucial information about cardiac rhythm, conduction abnormalities, and the overall health of the heart (1). Due to its non-invasive nature and ability to quickly provide valuable diagnostic insights, ECG is widely utilized in various medical settings, including emergency departments, intensive care units, and outpatient clinics. Unfortunately, even with educational initiatives, physicians at every stage of training showed shortcomings in interpreting ECGs (2). Electrolyte abnormalities, specifically imbalances in potassium, sodium, and calcium levels, can have profound effects on cardiac function. These abnormalities can disrupt the normal electrical activity of the heart, leading to arrhythmias, conduction disturbances, and other potentially life-threatening complications. Additionally, they are encountered commonly in the elderly and intensive care patient populations (3). Besides their high prevalence and mortality rates, it was shown that prompt detection and management of electrolyte abnormalities are essential for decreasing mortality and adverse cardiac events in various patient groups (4; 5; 6; 7). Currently, only blood sample analysis offers electrolyte concentration values, but a non-invasive diagnostic method applicable at the point of care, at home, or even retrospectively with existing measurements, could provide insight into the relationship between arrhythmias, including sudden cardiac death, and changes in plasma electrolyte concentrations (8). In this respect, ECG has been utilized for a while due to the strong dependency of plasma electrolytes and ECG pattern changes (9). In addition to the usefulness and ease of use, the growing popularity of mobile monitoring with smart devices enhances the appeal of this diagnostic method, as exemplified by studies effectively using the ECG for estimating electrolyte levels. These advancements enable early identification and intervention, which could potentially prevent severe cardiac complications (10; 11; 12). Recent advances in artificial intelligence (AI) have revolutionized the field of medical imaging and signal processing, offering new opportunities for enhancing the diagnostic capabilities of ECG (13). AI models, utilizing deep learning algorithms and neural networks, can analyze ECG signals with remarkable accuracy and efficiency. By leveraging large datasets of ECG recordings, AI models can learn to recognize subtle patterns and deviations indicative of electrolyte abnormalities, enabling earlier detection and intervention (14). Despite the growing interest in AI-based approaches for ECG analysis, there is currently a paucity of systematic reviews or meta-analyses examining the role of AI in detecting electrolyte abnormalities on ECG. In international prospective register of systematic reviews (PROSPERO), a preliminary search revealed no current or ongoing review of the diagnostic accuracy of AI models for the detection of electrolyte abnormalities. Therefore, the current study aims to conduct a systematic review and meta-analysis to evaluate the diagnostic accuracy of AI models in detecting electrolyte abnormalities from ECG recordings. The primary objective is to assess the sensitivity, specificity, and overall diagnostic performance of AI-based algorithms. Secondary aims include exploring the potential impact of AI on clinical decision-making and patient outcomes.

### Research Questions

This review addresses the following questions:

- Which AI models have been developed for the detection of electrolyte imbalances?
- Which databases and pre-processing methods have been utilized?
- What is the diagnostic accuracy of AI models in detecting electrolyte imbalances?
- Can AI models for detecting electrolyte imbalances from ECGs be effectively implemented in hospital settings?

## Methods

This systematic review and meta-analysis were conducted according to the Preferred Reporting Items for Systematic Reviews and Meta-Analyses of Diagnostic Test Accuracy (PRISMA-DTA) statement as attached in Supplementary Table 1. The review protocol was registered in the International Prospective Register of Systematic Reviews (PROSPERO) database (CRD42024530195) before the data analysis began.

### Search Strategy

The electronic database search was conducted in PubMed, MEDLINE, Cochrane Library, Embase, Elton B. Stephens Company, Web of Science, and Cumulated Index to Nursing and Allied Health Literature, Google Scholar, Scopus, IEEE, Arxiv, Bioarxiv and, Medarxiv for articles reporting the diagnostic accuracy of AI-based models detecting electrolyte abnormalities up to October 1st, 2024. The search terms were “AI”, “artificial intelligence”, “machine learning”, “deep learning”, “computer intelligence”, “neural network”, “kalemi*”, “kalaemi*”, “electrol*”, “potass*”, “calcium”, “calcemi*”, “sodium”, “natremi*”, “electrocardiography”, “ECG”, “electrocardi*”, “EKG”, and “cardiac electrophysiology”. The full search terms for each database are presented in Supplementary Table 2. Additional primary sources were acquired through a comprehensive approach, including manual searches of review studies and original articles to track references. Only original data were employed in the collection process of the meta-analysis.

### Eligibility Criteria and Study Selection

The inclusion criteria were as follows:

- Studies involving participants aged 15 years and older
- Studies reporting the diagnostic accuracy of deep learning or conventional machine learning-based AI models predicting electrolyte imbalances from ECG
- Studies having reference standards as health-care professionals or serum analysis-based diagnosis or final diagnosis for cases extracted from databases
- Studies having diagnostic accuracy measures including sensitivity, specificity, true positive, false positive, true negative, false negative
- Any type of diagnostic test accuracy studies including randomized controlled trials, prospective cohort studies, retrospective cohort studies, case-control studies
- Published or preprint studies
- Studies in English

The exclusion criteria were as follows:

- Non-human studies (animal, in-vivo, etc.) and studies covering pediatric population (<15 years of age)
- Studies utilizing simple algorithms without machine learning
- Studies lacking the quantitative performance metrics for the model
- Commentaries, editorials, abstracts without full-text articles, letters, expert opinions, systematic reviews, meta-analyses, and surveys
- Studies written in languages other than English
- Synthetic or simulated data use

No exclusion was made based on the date of publication. For the current study, we only included studies that used machine learning methods either in feature extraction or classification. All search results were exported to Mendeley Reference Manager (v2.113). Duplicates were eliminated, followed by the screening of titles and abstracts of all search results and included based on the eligibility criteria. Full-text articles were then obtained and independently assessed for final inclusion by two investigators. Any discrepancies were resolved through discussion with a third reviewer. Interrater reliability was assessed using Kappa statistics (κ) (15), with values interpreted as follows: 0, indicating no agreement; 0.01–0.20, signifying none to slight agreement; 0.21–0.40, indicating fair agreement; 0.41–0.60, suggesting moderate agreement; 0.61–0.80, representing substantial agreement; and 0.81–1.00, indicating almost perfect agreement (16).

### Assessment for Risk of Bias and Study Quality

Two reviewers independently evaluated the quality of the findings by using the Quality Assessment of Diagnostic Accuracy Studies-2 (QUADAS-2) scale which contains nine signaling questions under four domains for risk of bias (patient selection, reference standard, index test, and flow and timing) and three domains for applicability (patient selection, index test, and reference test) (17). Domains under risk of bias are classified as low risk if all items within that domain were assessed as “yes” and classified as high risk if any item was assessed as “no” (17). Any discrepancies were resolved through mutual discussion and consensus with a third reviewer. Weights were not assigned to the QUADAS item or summary score in the analysis, as conclusions regarding the impact of quality on estimates of diagnostic accuracy varied and were influenced by the methods used to generate the quality score (18). Therefore, subgroup analysis was conducted to investigate whether the scores on the quality items, including patient selection, index test, reference standard, and flow and timing, explained variations in diagnostic performance.

### Data Extraction

Two independent reviewers utilized a standardized form adapted from the JBI Reviewers’ Manual 2015 to extract data (19). Information extracted from each study included publication year, author, study design, data source, type of detection, algorithm type, index test, reference test, and model performance metrics. Pilot testing was conducted on five studies to ensure meaningful and efficient data extraction before proceeding with the remaining studies (20). Test performance data were extracted in the form of a 2×2 table comprising true positive, true negative, false positive, and false negative values directly from tabulated results. If these data were unavailable, they were derived from reported sensitivity, specificity, positive predictive value, and/or negative predictive values. Studies unable to undergo this calculation or those where doubt arose regarding the 2×2 calculation were excluded from subsequent analysis.

### Data Analysis

To assess the effectiveness of AI based electrolyte imbalance predictions from ECG, we conducted a metaanalysis. Among the electrolyte imbalances we evaluated, only hyperkalemia and hypokalemia were eligible to conduct meta-analysis on diagnostic test accuracy. We included the studies which 2×2 matrix was reported or can be calculated. For meta-analysis, hyperkalemia was defined as serum potassium level>5.5 mmol/L, hypokalemia was defined as serum potassium level<3.5 mmol/L (21). Studies or results that evaluated diagnostic accuracy using different threshold values for hyperkalemia (e.g., >5 or >6 mmol/L) or hypokalemia (e.g., <3 or <2.5 mmol/L) were excluded from the meta-analysis. The meta-analyses were conducted using R version 4.3.3 software and the mada, metafor and meta packages (22; 23; 24). Pooled specificity, sensitivity, Positive Likelihood Ratio (PLR), Negative Likelihood Ratio (NLR), and diagnostic odds ratio (DOR) were estimated by integrating individual studies and were summarized using the forest plot with 95% confidence intervals. The Clopper–Pearson method was used as the default approach to calculate confidence intervals (23). For proportion-type data, logit transformation was applied. Accuracy estimates were pooled using a random-effects model and between study variance was estimated by the DerSimonian-Laird method. Study heterogeneity was quantified using Higgin’s I^2^ and Cochrane’s Q statistics. To summarize diagnostic performance, a summary receiver operating characteristic (SROC) curve was created using a bivariate model. Additionally, meta-regression was conducted to investigate the effects of potential moderators (e.g., number of leads, testing environment, region, patient selection, and model type).

## Results

### Study Characteristics

We conducted a comprehensive search across nine different databases, resulting in 763 articles after removing duplicates (Figure 1A). Screening these articles by title and abstract identified 40 studies as relevant to our systematic review. A further assessment of eligibility narrowed this selection to 21 studies included in the systematic review (25; 26; 27; 28; 11; 29; 30; 31; 32; 33; 34; 35; 36; 37; 38; 39; 40; 41; 42; 43; 44) (Table 1, Figure 1B). Of these, only 10 were suitable for inclusion in the meta-analysis (25; 11; 29; 30; 31; 32; 33; 34; 36; 39; 41; 42; 43; 44) (Table 1, Figure 1B). Inter-rater reliability between two reviewers for data selection was 0.821. During the screening process, we assessed the internal and external validation testing metrics of the developed models separately. This distinction is crucial in machine learning. Deep learning models often perform well on datasets similar to those on which they have been trained. However, testing these models outside their training environment presents significant challenges. Variations in data collection methods, devices used, and operational teams can introduce subtle changes to the dataset, a phenomenon known as the out-of-domain problem in machine learning (45). Our review specifically aimed to evaluate how AI models designed for detecting electrolyte imbalances performed under both internal and external validation conditions. It’s important to note that our analysis focused solely on the models’ performance on test datasets, without considering their training data results. Additionally, personalized and dynamic model development was excluded from the meta-analysis (27; 37). This exclusion is crucial because such models may not be generalized for diagnostic purposes, and comparing their performance with other studies may not be fair due to the potential overestimation of their effectiveness. Data used in the studies originated from five countries: China, South Korea, Sweden, Taiwan, and the USA (Table 1, Supplementary data). Most of the studies collected their own datasets and primarily focused on ECG-based predictions. However, Chiu et al. and Babur et. al. (26; 27) utilized the publicly available MIMIC-III dataset to classify heartbeats and diseases respectively. One study did not disclose its data source (31). The total number of test ECG recordings varied widely, ranging from 32 to 130,145. Cumulatively, a total of 671,062 ECGs were tested across all studies, although a single ECG might have been used in multiple studies or for different electrolyte predictions. Apart from that, Chiu IM et al.’s study contributed 105,546 heartbeats from 278 patients. A breakdown of ECG recordings used for testing includes 222,027 from Taiwan (34; 35; 37; 38; 43), 122,609 from Sweden (40), 124,014 from the USA (28; 11; 29; 30), 198,273 from South Korea (25; 32; 33; 39), and 3761 from China (41; 42; 44). Of these, 298,848 recordings were used for internal testing and 372,214 for external testing. The largest external testing was conducted by An et. al. with test size of 130,145 ECGs (25). Notably, Galloway et al. tested their deep learning model on three different domains from Minnesota, Florida, and Arizona, with test sizes of 50,009, 6011, and 5855, respectively (11)(Supplementary data). ECGs are typically recorded using 12 leads. However, among the 21 studies evaluated, configurations varied: 11 studies tested models using 12 leads, 7 employed a single lead (Lead II), three utilized 2 leads, two used 4 leads, one used 3 leads, one used 6 leads, one used 7 leads, and another used 8 leads. Notably, Galloway et. al. (11) experimented with both 2 leads (Leads I and II) and 4 leads (Leads I, II, V3, and V5). An et. al. (25) tested on 12, 3 and single leads. Kwon et al. (33) explored configurations of 12, 6, and 1 lead, while Wang et al. (41) focused on 12 leads and a single lead (Table 1, Supplementary data). Some studies applied feature extraction methods to train their models, whereas others utilized raw ECG data. Most of the studies processed the ECG data as signals, with the exceptions of Wu et al. (43), Wang et al. (41) and Kim et. al. (32) who processed the ECGs as image data. We classified the models used in the studies under two categories as traditional machine learning algorithms and deep learning algorithms. Among 21 studies, one study used only support vector machine (SVM) for classification of hyperkalemia (31) and the rest of the studies employed at least one deep learning algorithms. Most of the deep learning algorithms used in the studies were CNN based algorithms. Xu et. al. (44) experimented both CNN based algorithm and machine learning algorithms including SVM, extreme gradient boosting (XGB), adaptive boosting (AdaBoost) and logistic regression for detection of hyperkalemia (Table 1). Several studies have developed bespoke machine learning or deep learning models for electrolyte imbalances (Table 1, Figure 1B). Kwon et al. (33) developed a CNN model using 83,449 ECGs to independently predict potassium, calcium, and sodium levels. Khan et al. (31) focused on the classification of six different cardiac conditions, including hyperkalemia and hypercalcemia. Von Bachmann et al. (40) created a deep learning-based regression model using eight leads of ECG (I, II, V1-V6) to predict serum levels of potassium, calcium, and sodium. Similarly, Babur et. al. (26) used neural network-based regression to predict serum potassium and calcium levels. Lou et al. (37) developed ECG embeddings with a deep learning model and used these embeddings to predict various patient characteristics, including electrolyte levels (K, Ca, Na).

**Table 1.**
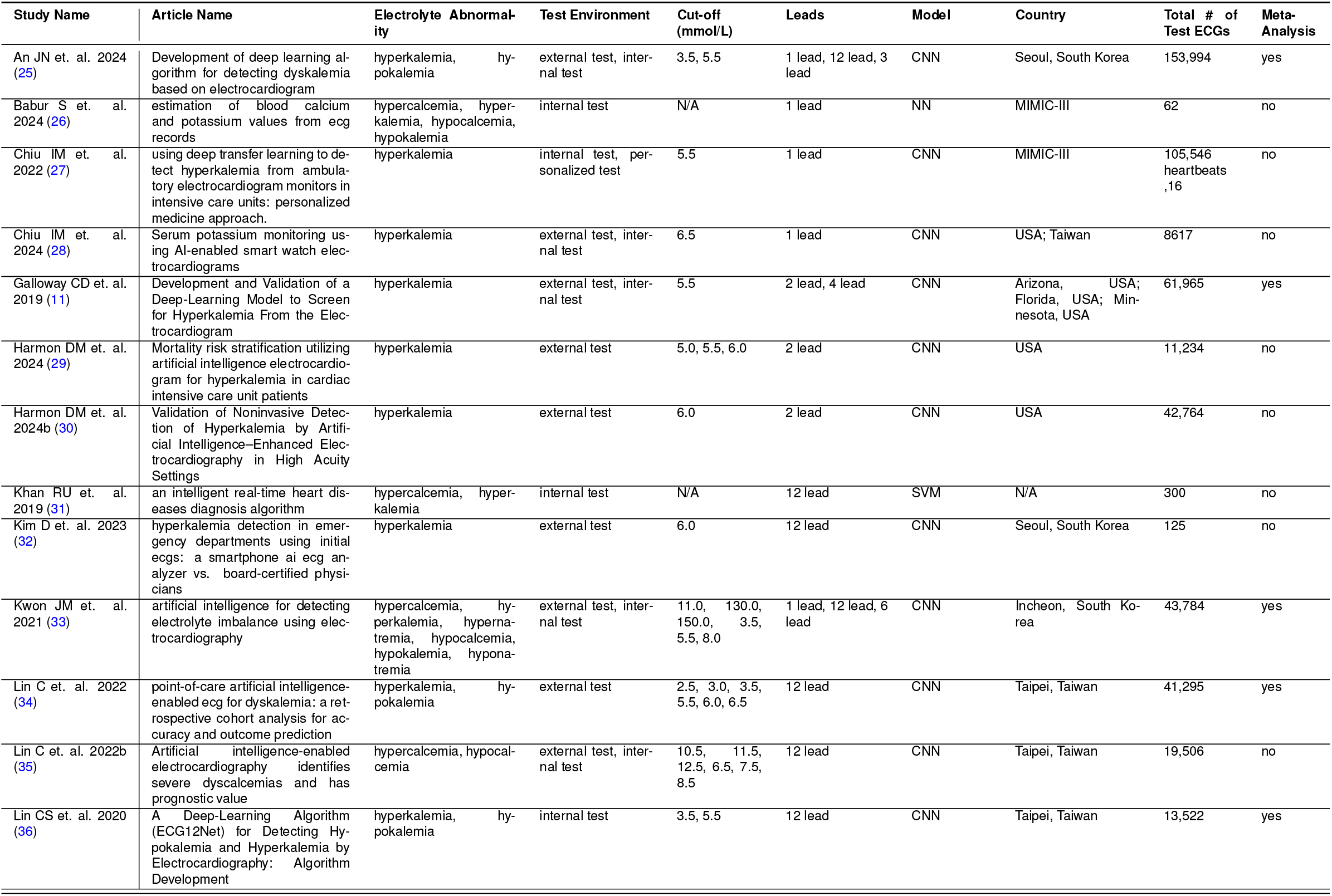
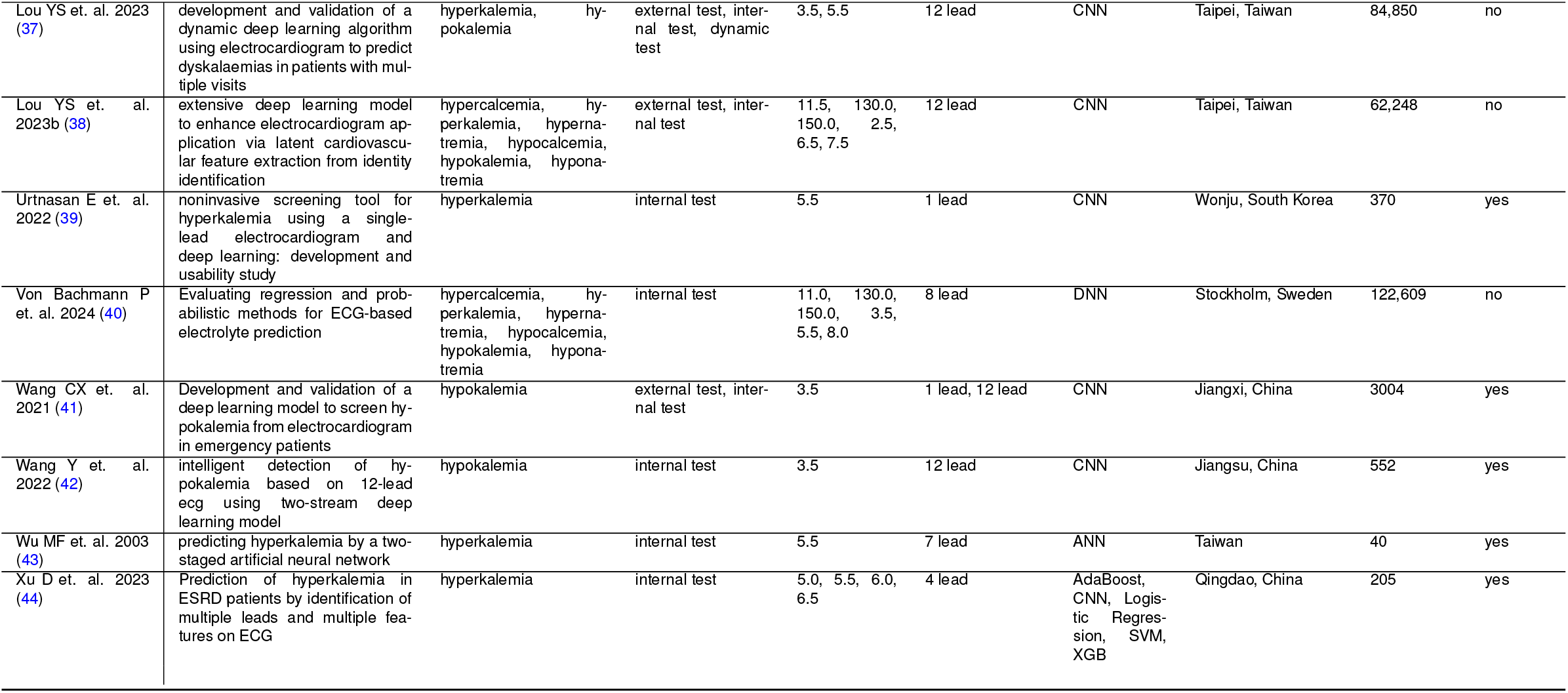
Studies included in the systematic review.

**Figure 1.**
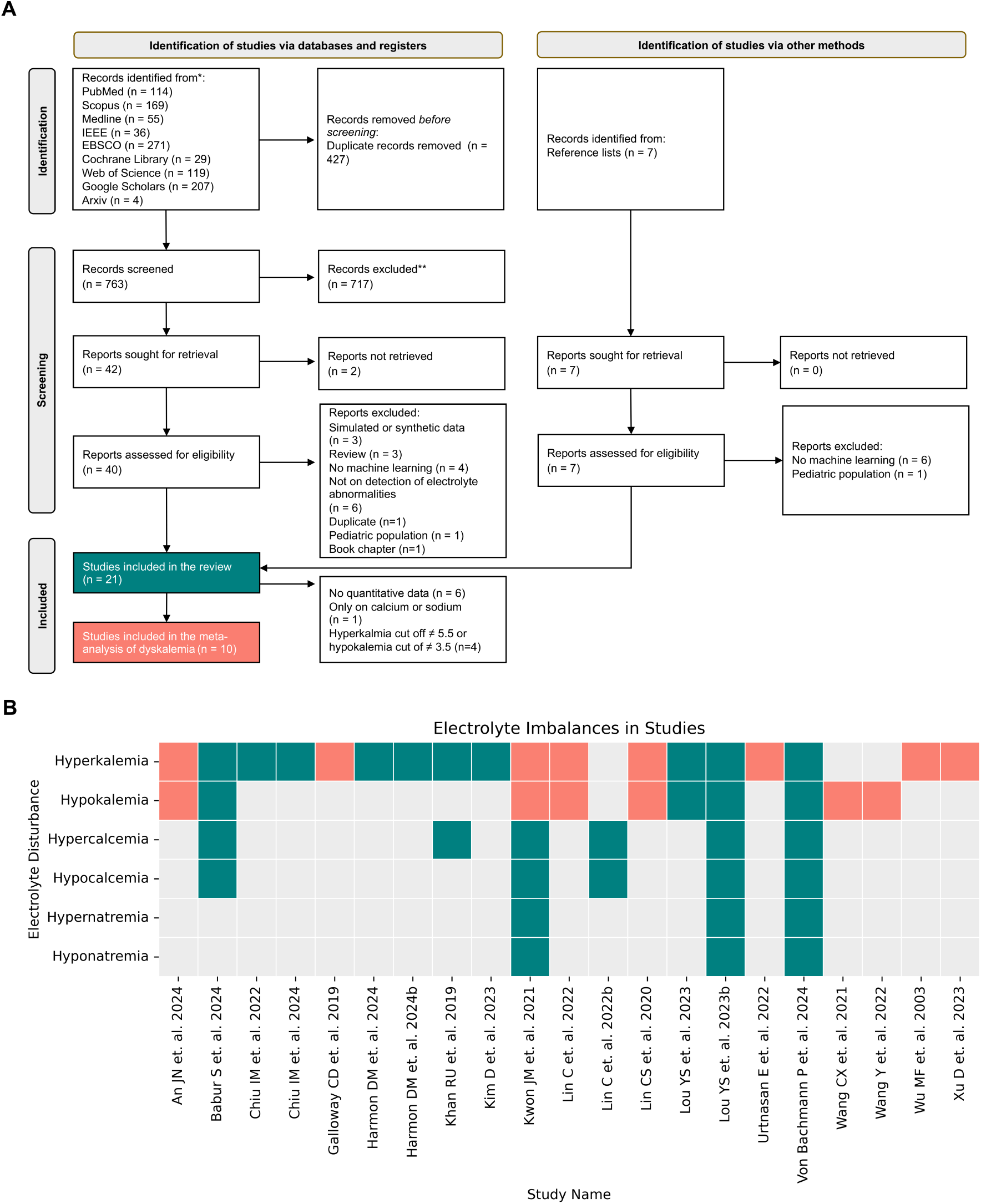
PRISMA 2020 flow chart. **(A)** A total of 21 studies were included in the systematic review, 10 of which were also included in the meta-analysis. **(B)** Studies included in the systematic review are represented alongside the electrolyte abnormalities they examine, with each study depicted as a colored square. Orange squares indicate studies that were additionally included in the meta-analysis on hyperkalemia and hypokalemia.

### Assessment for risk of bias

The results of QUADAS-2 for studies that are included in the review are illustrated in Figure 2. Inter-rater reliability and detailed evaluations for each study are provided in Supplementary Table 3, 4. More than %50 of the studies (11/21) displayed high bias in patient selection, mainly due to lack of information on sampling methods and avoidance of case-control approach. Therefore, we conducted a sub-analysis focusing on patient selection bias in the meta-analysis. Concerning the index test, 17 out of 21 studies (81%) reported low risk of bias whereas 2 had high, 2 had unclear bias. These 4 studies had vague information on whether the index test results were interpreted without knowledge of the results of the reference standard and whether the threshold was prespecified. Ideally, multiple ECGs from the same patient should not be split between the training and test sets, as this could lead to an overestimation of performance. 4 study had unclear bias in the reference standard domain, failing to explain how the reference standard, serum electrolyte level measurement, was conducted. In terms of flow and timing, 11 studies were rated as having a high or unclear risk of bias, either because not all patients were included in the analysis or due to an inappropriate interval between the index test and the reference standard. For potassium, we considered an interval of 2 hours appropriate, while for calcium, the acceptable interval was 4 hours.

**Figure 2.**
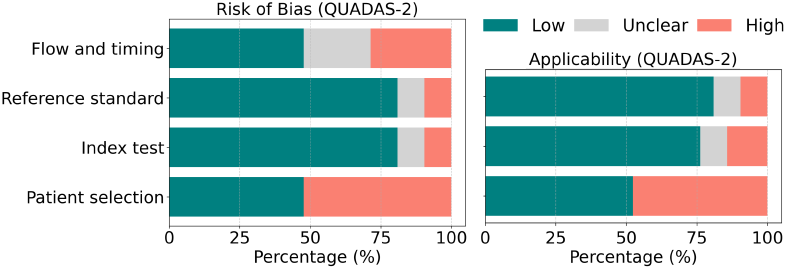
Graphical display for QUADAS-2 results. QUADAS-2 domains show proportion of studies with low, high or unclear risk of bias and applicability concerns.

### Potassium Imbalance

Most of the studies focused on potassium imbalances (n = 20), with 18 addressing hyperkalemia and 10 examining hypokalemia (Figure 1B, Table 1). This focus is reasonable considering the life-threatening clinical significance of hyperkalemia and its detectability on ECGs due to prominent features. 12 studies utilized external datasets to validate their models for hyperkalemia and hypokalemia detection. Among the studies focusing on potassium imbalances, six applied preprocessing algorithms to extract features from ECG signals. Khan et al. (31) utilized morphological operations to extract features such as P, Q, R, S, and T waves, including the width and height of each wave, from an ECG signal. They also removed non-relevant time areas using discrete wavelet transform. One of the earlier studies (43) extracted five features from lead II, including P wave duration, RR interval, corrected QT interval, and QRS complex width. Additionally, they extracted twelve more features, such as the amplitude and duration of T waves across the V1-V6 chest leads, for the second stage of their neural network. Xu D. et al. (44) extracted twelve different ECG characteristics, including T right slope, T left slope, S-T band slope, T wave amplitude, R wave amplitude, S wave amplitude, T wave area, R wave area, S wave area, and the areas per second of T, R, and S waves from leads V2-V5, totaling 48 ECG features. Babur et. al. (26) extracted features related to time, frequency and time-frequency domains in addition to morphological features. Wang et al. (42) employed the Pan-Tompkins algorithm to locate the QRS complex and then extracted the following six features: the proportion of T-wave width to the whole RR interval, peak-to-peak value of the T-wave, slope between the start of the T-wave and the first peak, slope between the end of the T-wave and the second peak, integral value between the start and end of the T-wave, and the ratio of the distance between the peak of the R-wave and the end of the T-wave to the RR interval. They proposed concatenating these calculations with CNN output, achieving better results than using only CNN classification. Similarly, Lin et al. (36) included eight morphological ECG features—heart rate, PR interval, QRS duration, QT interval, QTc, P wave axis, RS wave axis, and T wave axis—in their analysis and concatenated them with the output of the CNN-based ECG12Net model to predict serum potassium levels. However, they observed that adding morphological features did not improve the results. Therefore, we included only the performance of the CNN-based model from Lin C.S. et. al.’s study in our meta-analysis to avoid duplication. Some studies reported model performance across various potassium thresholds or different severities of serum potassium levels, as detailed in Table 1 and Supplementary data. In our meta-analysis for hyperkalemia detection, we used a cutoff point of mmol/L, and for hypokalemia, a cutoff point of 3.5 mmol/L. For studies with multiple architectures, electrolytes, cutoff points, or testing environments, we provided detailed descriptions, with sub-study names indicated in parentheses next to the study name (e.g., (S1), (S2), etc.) (Supplementary data). Our meta-analysis included a total of 41 entities derived from 8 studies focusing on the detection of hyperkalemia (Figure 3). These entities varied based on internal versus external dataset evaluations, model architectures, and the number of ECG leads used. The pooled sensitivity and specificity for detecting hyperkalemia were 0.856 (95% CI: 0.829-0.879) and 0.688 (95% CI: 0.744-0.826), respectively. The diagnostic odds ratio (DOR) was 21.8 (95% CI: 17.8-26.7) (Table2, Supplementary Figure 1). It corresponded with a positive likelihood ratio (PLR) of 3.98 (95% CI:3.52-4.50) and a negative likelihood ratio (NLR) of 0.19 (95% CI: 0.17-0.22) (Supplementary Figure 2). In terms of dataset source, internal datasets showed a higher specificity 0.826 (95% CI: 0.722-0.869) compared to external datasets 0.738 (95% CI:0.672-0.795), although external datasets had slightly better sensitivity 0.864 (95% CI: 0.840-0.885) (Table 2). Regionally, studies from Asia exhibited a balanced performance with a sensitivity of 0.852 (95% CI: 0.814-0.883) and specificity of 0.817 (95% CI: 0.765-0.859). The analysis by the number of leads used showed that using 12 leads yielded DOR of 35.6 (95% CI: 26.5-47.9), indicating a robust model performance. Notably, single-lead setups demonstrated a higher pooled sensitivity but lower pooled specificity than 12 leads configuration. For all 41 different entities, the heterogeneity (I2) was 95% (sensitivity), 100% (specificity), and 91% (DOR). According to summary receiving operating characteristics curve (SROC), area under the curve (AUC) was 0.891 (Figure 5A). In the case of hypokalemia, our meta-analysis included 27 entities from 6 studies (Figure 4). The pooled sensitivity was 0.824 (95% CI: 0.785-0.856), and the pooled specificity was 0.724 (95% CI: 0.668-0.774). The DOR was relatively lower at 12.27 (95% CI:9.15-16.47) (Table 2, Supplementary Figure 3). This analysis corresponded with a PLR of 3.02 (95% CI: 2.53-3.61) and an NLR of 0.25 (95% CI: 0.21-0.29) (Supplementary Figure 4). Subgroup analysis revealed that testing on internal datasets had higher sensitivity and specificity compared to external dataset. When analyzed by the number of leads, 12-lead configurations showed a modest sensitivity of 0.797 (95% CI: 0.731-0.851) and a specificity of 0.764 (95% CI: 0.698-0.819). Notably, six-lead setups displayed the highest sensitivity at 0.907 (95% CI: 0.893-0.919) but with lower specificity. For all 27 different entities, the heterogeneity (I2) was 99% (sensitivity), 100% (specificity), and 99% (DOR). According to SROC analysis, AUC was 0.846 (Figure 5B). Deek’s funnel plot showed no publication bias for both hyperkalemia and hypokalemia (Supplementary Figure 5, p values are 1.0 and 0.191, respectively)

**Table 2.** Subgroup Analysis for Hyperkalemia and Hypokalemia. * indicates significant difference compared to reference within each subgroup. The first study types within each subgroup are considered the reference group. Detailed meta-regression results are in Supplementary Table 5-10.

| Subgroup | Number of entities | Pooled sensitivity [95% CI] | Pooled specificity [95% CI] | Pooled DOR [95% CI] |
| --- | --- | --- | --- | --- |
| **Hyperkalemia** |  |  |  |  |
| All studies | 41 | 0.856 [0.829-0.879] | 0.788 [0.744-0.826] | 21.80 [17.80-26.69] |
| *Dataset* |  |  |  |  |
| Internal | 22 | 0.845 [0.794-0.886] | 0.826 [0.772-0.869] | 25.76 [18.84-35.22] |
| External | 19 | 0.864 [0.840-0.885] | 0.738 [0.672-0.795] | 18.39 [14.20-23.81]* |
| *Region* |  |  |  |  |
| Asia | 29 | 0.852 [0.814-0.883] | 0.817 [0.765-0.859] | 25.56 [19.31-33.85] |
| USA | 12 | 0.863 [0.832-0.890] | 0.707 [0.654-0.756]* | 15.64 [13.47-18.17]* |
| *Number of leads* |  |  |  |  |
| 12 leads | 10 | 0.849 [0.779-0.899] | 0.867 [0.787-0.920] | 35.61 [26.48-47.87] |
| 7 leads | 1 | 0.600 [0.381-0.786] | 0.650 [0.426-0.823]* | 2.79 [0.77-10.04]* |
| 6 leads | 2 | 0.903 [0.872-0.928] | 0.716 [0.505-0.862] | 23.37 [5.07-107.72] |
| 4 leads | 11 | 0.835 [0.772-0.884] | 0.771 [0.705-0.826]* | 18.88 [16.70-21.35]* |
| 3 leads | 4 | 0.868 [0.807-0.911] | 0.796 [0.707-0.863] | 26.15 [23.89-28.63] |
| 2 leads | 6 | 0.850 [0.799-0.890] | 0.690 [0.606-0.763]* | 13.15 [10.87-15.90]* |
| 1 lead | 7 | 0.887 [0.830-0.927] | 0.776 [0.635-0.873]* | 27.69 [10.14-75.59]* |
| *Patient selection bias* |  |  |  |  |
| Low | 10 | 0.856 [0.788-0.905] | 0.838 [0.711-0.915] | 30.54 [17.56-53.12] |
| High | 31 | 0.856 [0.827-0.881] | 0.769 [0.729-0.805] | 19.68 [16.63-23.30] |
| *Model type* |  |  |  |  |
| Machine learning | 4 | 0.695 [0.615-0.764] | 0.860 [0.801-0.904] | 14.45 [9.93-21.02] |
| Deep learning | 37 | 0.866 [0.842-0.887]* | 0.779 [0.730-0.820] | 22.63 [18.16-28.20] |
| *Sample size* |  |  |  |  |
| ≤1000 | 8 | 0.792 [0.642-0.890] | 0.869 [0.768-0.929] | 24.99 [8.53-73.23] |
| >1000 | 33 | 0.866 [0.844-0.886] | 0.767 [0.720-0.808]* | 21.42 [17.91-25.62]* |
| **Hypokalemia** |  |  |  |  |
| All studies | 27 | 0.824 [0.785-0.856] | 0.724 [0.668-0.774] | 12.27 [9.15-16.47] |
| *Dataset* |  |  |  |  |
| Internal | 14 | 0.842 [0.782-0.887] | 0.737 [0.672-0.793] | 14.98 [9.90-22.68] |
| External | 13 | 0.802 [0.759-0.840] | 0.709 [0.612-0.789] | 9.95 [6.65-14.89] |
| *Number of leads* |  |  |  |  |
| 12 leads | 13 | 0.797 [0.731-0.851] | 0.764 [0.699-0.819] | 12.79 [8.11-20.17] |
| 6 leads | 2 | 0.907 [0.893-0.919] | 0.543 [0.394-0.684] | 11.40 [6.00-21.63] |
| 3 leads | 4 | 0.823 [0.749-0.879] | 0.806 [0.755-0.849] | 19.14 [16.11-22.73] |
| 1 lead | 8 | 0.838 [0.774-0.886] | 0.642 [0.526-0.744]* | 9.30 [5.00-17.32] |
| *Patient selection bias* |  |  |  |  |
| Low | 10 | 0.839 [0.760-0.895] | 0.643 [0.521-0.748] | 9.21 [6.27-13.53] |
| High | 17 | 0.814 [0.774-0.849] | 0.766 [0.719-0.808]* | 14.39 [9.73-21.28] |
| *Sample size* |  |  |  |  |
| ≤1000 | 2 | 0.736 [0.662-0.799] | 0.861 [0.671-0.950] | 16.34 [5.74-46.52] |
| >1000 | 25 | 0.829 [0.791-0.862] | 0.711 [0.655-0.762] | 12.02 [8.82-16.38] |

**Figure 3.**
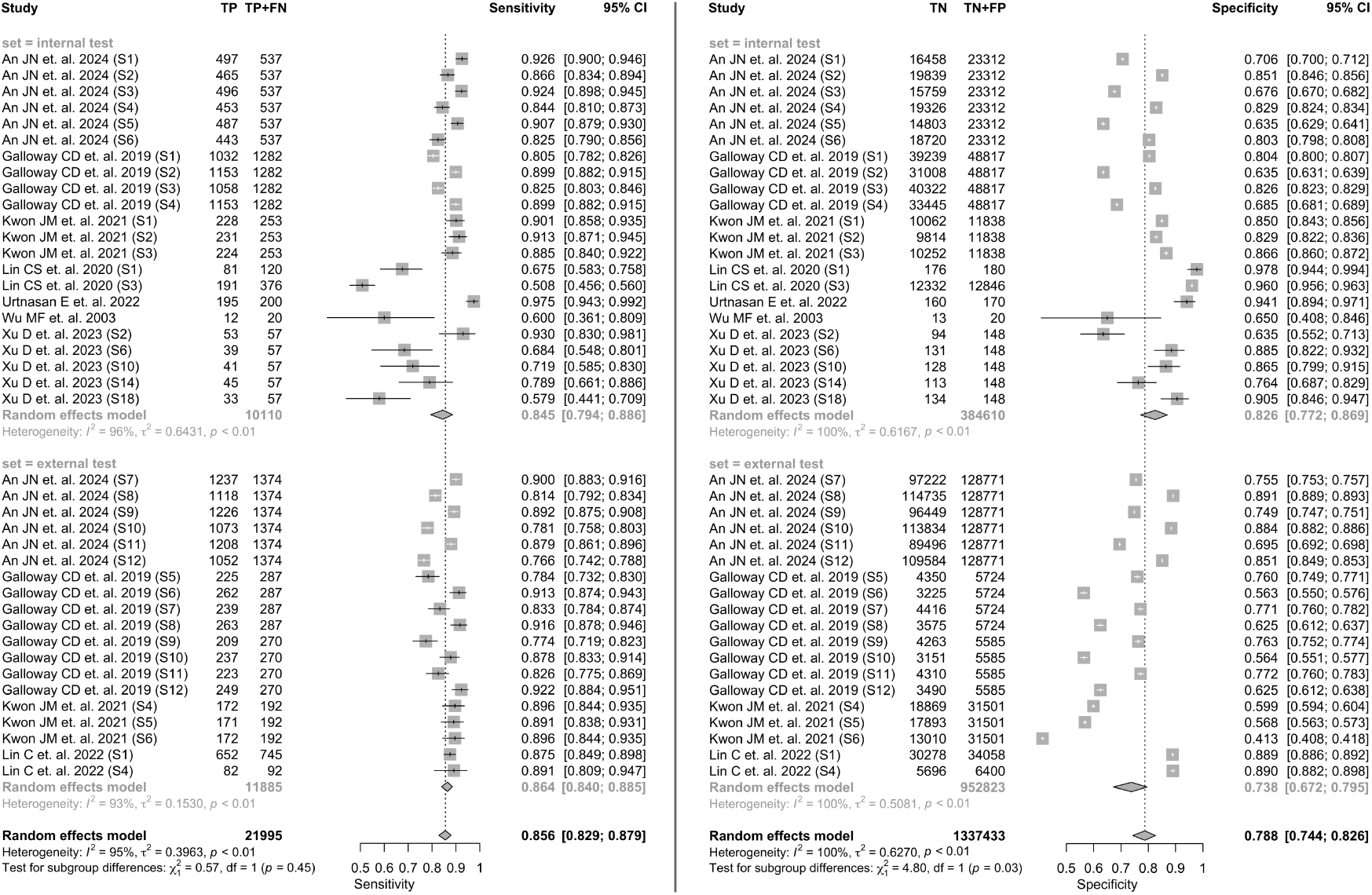
Forest plots of sensitivity and specificity of included studies for hyperkalemia. The pooled sensitivity and specificity of AI models in hyperkalemia detection are 0.856 and 0.788, respectively.

**Figure 4.**
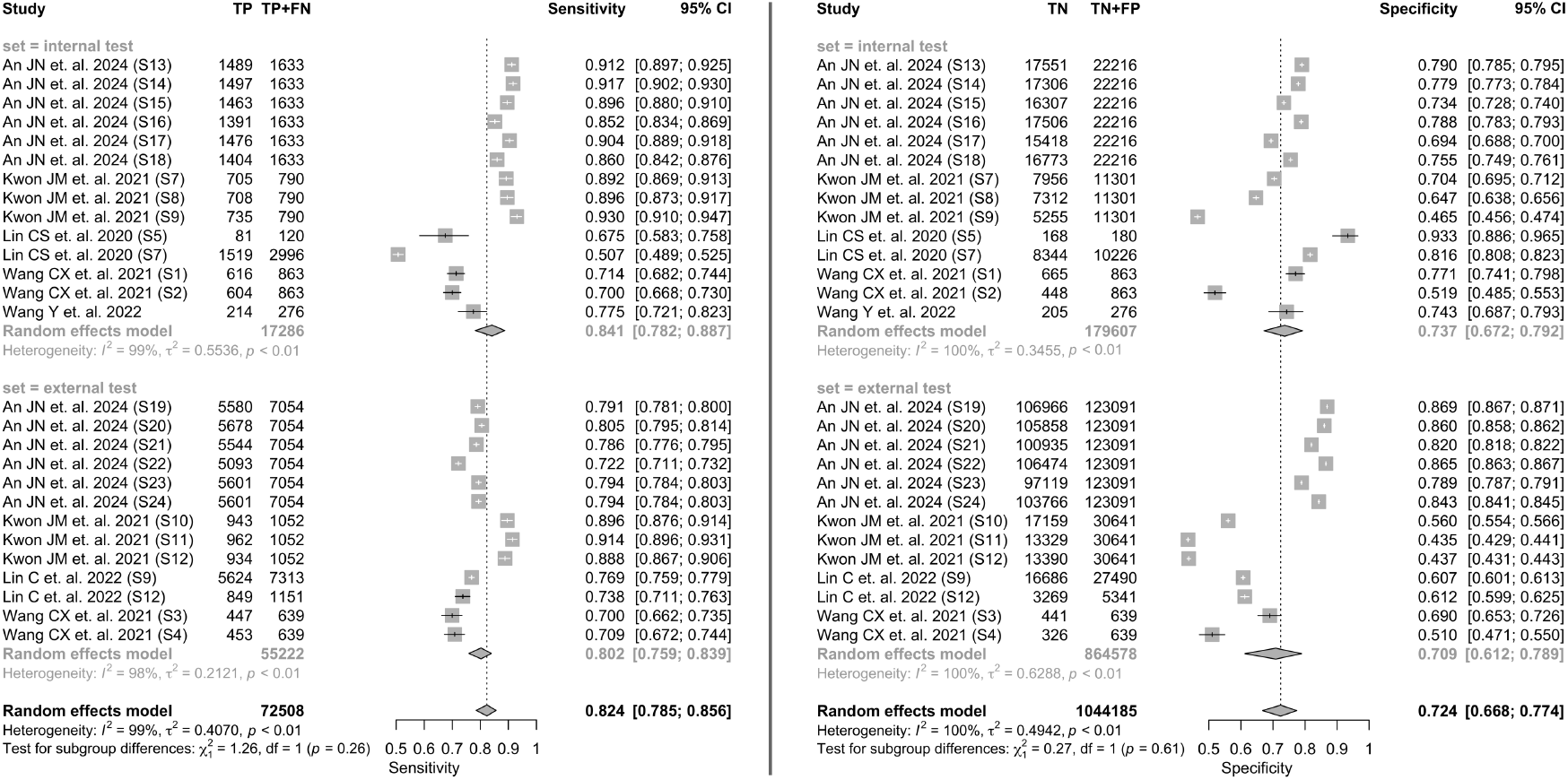
Forest plots of sensitivity and specificity of included studies for hypokalemia. The pooled sensitivity and specificity of AI models in hypokalemia detection are 0.824 and 0.724, respectively.

**Figure 5.**
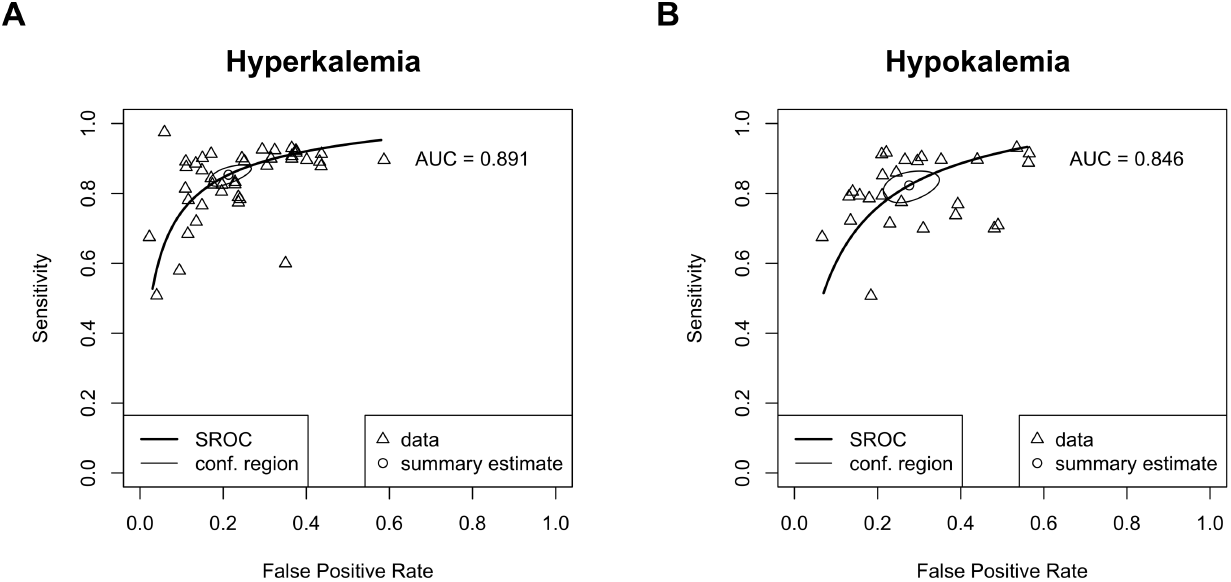
Summary receiver operating characteristic (SROC) curves for hyperkalemia and hypokalemia studies. The area under the curve (AUC) is 0.891 for hyperkalemia and 0.846 for hypokalemia. Triangles indicate individual entities included in the meta-analysis, while the circle represents the summary estimate of AI model performance.

### Calcium Imbalance

There were six studies on the detection of calcium imbalances, including two from Taiwan (35; 37), one from South Korea (Kwon et al., 2021), and one from Sweden (40). Babur et. al. (26) used MIMIC-III dataset (Table 1, Supplementary data). These studies focused on detecting both hypercalcemia and hypocalcemia. Khan R.U. et. al. (31) did not specify the data source or study environment. All studies utilized deep learning models except for R.U. Khan et. al. Kwon J.M. et al. developed a convolutional deep learning model using 83,449 ECGs, experimenting with different numbers of leads. They conducted both internal and external validation tests. Lin C. et al. employed the CNN-based ECG12Net architecture, which had previously been used for hyperkalemia and hypokalemia detection. They demonstrated that detection performance increases with the severity of calcium imbalance. The model’s AUC in external test data was 0.845 for severe hypercalcemia and 0.772 for severe hypocalcemia, respectively. Babur et. al. used neural network-based regression for estimating serum calcium levels.

### Sodium Imbalance

Three articles investigated sodium imbalance detection from ECGs using AI (33; 37; 40) (Table 1, Supplementary data). Kwon J.M. et.al experimented developing deep learning model on single, 6-lead, and 12-lead ECGs for sodium imbalance. Internal validation showed sensitivities of 0.846 for 1 lead and 0.923 for both 6 and 12 leads in detecting hypernatremia. The specificities ranged from 0.347 for a single lead to 0.634 for 12 leads. External validation revealed sensitivities of 0.907 for 1 lead, 0.889 for 6 leads, and 0.870 for 12 leads, with specificities increasing with the number of leads, from 0.253 for a single lead up to 0.649 for 12 leads. For the diagnosis of hyponatremia, the internal validation for detecting hyponatremia demonstrated sensitivities of 0.912 for 1 lead, 0.89 for 6 leads, and 0.901 for 12 leads. The specificities were 0.686 for a single lead, 0.797 for 6 leads, and 0.82 for 12 leads. In external validation, sensitivities were 0.915 for 1 lead, 0.887 for both 6 and 12 leads, while specificities were 0.477 for a single lead, 0.599 for 6 leads, and 0.629 for 12 leads (Supplementary data). In Lou Y.S. et. al.’s study, for hypernatremia, sensitivity, specificity and AUC was 0.431, 0.914 and 0.684 in external validation, respectively. For hyponatremia, they found sensitivity of 0.641, specificity of 0.686 and AUC of 0.704 in external dataset testing. Von Bachmann P. et. al. has showed the superior performance of DNN based regression over classical regression models in detection of sodium imbalance.

### Interpretability of models

Chiu et. al. (27) applied the Grad-CAM algorithm to a CNN model, demonstrating that the QRS complex was associated with the detection of hyperkalemia. Similarly, Kwon et al. (33) utilized a gradient class activation map as a sensitivity map with the gradient backpropagation method, showing that their deep learning model focused on the QRS complex for detecting hyperkalemia, hypokalemia, and hyponatremia. Additionally, their model emphasized the T wave for hyperkalemia detection and the S wave for identifying hypernatremia and hypercalcemia. Lin et. al. (35) employed an XGBoost (XGB) classifier on a tuning set to identify key ECG features for dyscalcemia detection. Their deep learning model (DLM) revealed that ECG patterns associated with hypercalcemia included a shorter QT interval, prolonged PR interval, higher T wave axis, lower RS wave axis, prolonged QRS duration, increased heart rate, and a higher prevalence of left ventricular hypertrophy compared to normal ECG cases. Conversely, ECG patterns indicative of hypocalcemia included a prolonged QT interval, shorter PR interval, smaller T wave axis, higher RS wave axis, prolonged QRS duration, and a slightly increased heart rate. Notably, the explainable variation due to these known ECG features accounted for only 17.93% and 10.96% in internal and external validation sets, respectively, suggesting the presence of additional features identified by the DLM. Both Lin et al. (34) and Lin CS et al. (36) utilized class activation mappings in CNN models. Lin CS demonstrated that their deep learning model recognized ST segment depression in hypokalemia, as well as tented T waves and wide QRS complexes in hyperkalemia. An et. al. (25) adopted a feature-masking approach, directly obscuring key ECG features to evaluate their importance in detecting dyskalemia. They found that masking the P wave had minimal impact on detecting hypokalemia and hyperkalemia. However, masking the QRS and T waves significantly affected the model’s performance.

## Discussion

The principal findings of this systematic review were that the diagnostic accuracy of AI-based models for the detection of hyperkalemia was moderate with sensitivity and specificity of 0.856 (95% CI: 0.829-0.879) and 0.788 (95% CI: 0.744-0.826) respectively. The overall pipeline of studies aimed at detecting electrolyte abnormalities or predicting serum electrolyte levels is summarized in Figure 6A. Applying the point estimates of sensitivity and specificity for AI-based detection of hyperkalemia to a hypothetical cohort of 1000 individuals suspected of having hyperkalemia, with a prevalence rate of 10%—where 100 individuals actually have the disease—the AI models are likely to miss 14 cases and incorrectly identify 191 cases as diseased. Consequently, out of 276 cases identified as positive, 86 would be true positives and 191 false positives. Similarly, out of 724 cases identified as negative, 709 would be true negatives and 14 false negatives (Figure 6B). However, it should be notified that there were outliers in the meta-analysis that may underestimate the pooled performance. Our meta-analysis revealed significant heterogeneity across studies in testing procedures, data sources, data types, and algorithm types. According to subgroup analysis, external dataset validation had higher pooled sensitivity compared to internal validation. However, the pooled performance metrics comparing internal and external studies may not be entirely appropriate, as not all studies included both types of validation. In cases where studies conducted both types of testing, internal test metrics were typically higher than those for external testing. For example, in the model developed by Kwon et al. for hyperkalemia detection using a 12-lead ECG, the sensitivities were 90.1% for internal validation and 89.6% for external validation. Specificities were 85.0% for internal validation and 59.9% for external validation, with AUCs of 0.945 and 0.873, respectively. Notably, studies showed a greater disparity in specificity between internal and external validations. The diagnostic accuracy of AI-based detection of hyperkalemia demonstrated lower sensitivity but higher specificity and DOR in Asian studies compared to studies in the USA. However, it cannot be concluded that these differences are due to ethnographic factors, especially since only one study from the USA contributed to the meta-analysis. Future research should be conducted in various countries and regions to better understand the real impact of ethnic groups on the diagnostic performance of AI. Among the studies that investigated the effect of different number of leads, 12-lead configuration consistently deliver the highest AUC and overall balanced performance. This is possibly because 12-lead ECGs provide more comprehensive cardiac information, allowing for more precise analysis. 1-lead models are suitable for wearable devices or resource-limited environments but trade some accuracy for simplicity. The type of AI model employed significantly impacts diagnostic outcomes. Traditional machine learning models demonstrated a lower sensitivity (0.695) but higher specificity (0.860) compared to deep learning models, which showed higher sensitivity (0.866) but lower specificity (0.779). This difference might result from the type of input data used. Studies employing traditional machine learning algorithms mostly utilized hand-crafted features such as T wave amplitude, T wave duration, QT interval, etc., for detecting hyperkalemia, which may have limitations in detection accuracy. Indeed, a study by Xu D. et al. tested various algorithms, including deep learning and machine learning models, on 48 ECG features and found that XGB and SVM performed slightly better than CNN-based models. Studies that utilized the entire ECG generally achieved higher model performance, suggesting that additional useful information within the ECG could aid in identifying hyperkalemia. Deep learning algorithms may capture and model more complex patterns and relationships in large datasets, which might be more adept at recognizing varied presentations of hyperkalemia. Studies on calcium and sodium imbalances are notably limited compared to those on potassium. This discrepancy can be attributed to several factors. Unlike potassium, calcium and sodium imbalances generally have less pronounced effects on cardiac electrical activity and are less likely to produce specific or identifiable patterns on ECG (40). Potassium imbalances, particularly hyperkalemia, are well-known for their characteristic ECG changes, such as peaked T waves, QRS widening, and arrhythmias, which facilitate detection through AI models (46). In contrast, calcium and sodium abnormalities, while clinically significant, often manifest more subtly on ECG or lack consistent findings, making it more challenging for AI algorithms to detect these imbalances with high accuracy (46; 40). Moreover, the impact of these electrolytes on cardiac function may not be as immediately life-threatening or evident as severe dyskalemia, potentially reducing the urgency and focus on developing AI models for their detection. Using AI to check potassium levels from ECGs may not be the most cost-effective approach in hospital settings, where laboratory tests can provide immediate results. This is especially true given the current models’ sensitivity and specificity and the costs associated with setting up the necessary infrastructure. However, integrating AI systems into wearable devices could offer significant benefits for continuous monitoring of cardiac events and electrolyte disturbances from ECGs at home. This approach could be particularly valuable for elderly individuals with comorbidities, including renal diseases, enabling consistent and non-invasive health tracking.

**Figure 6.**
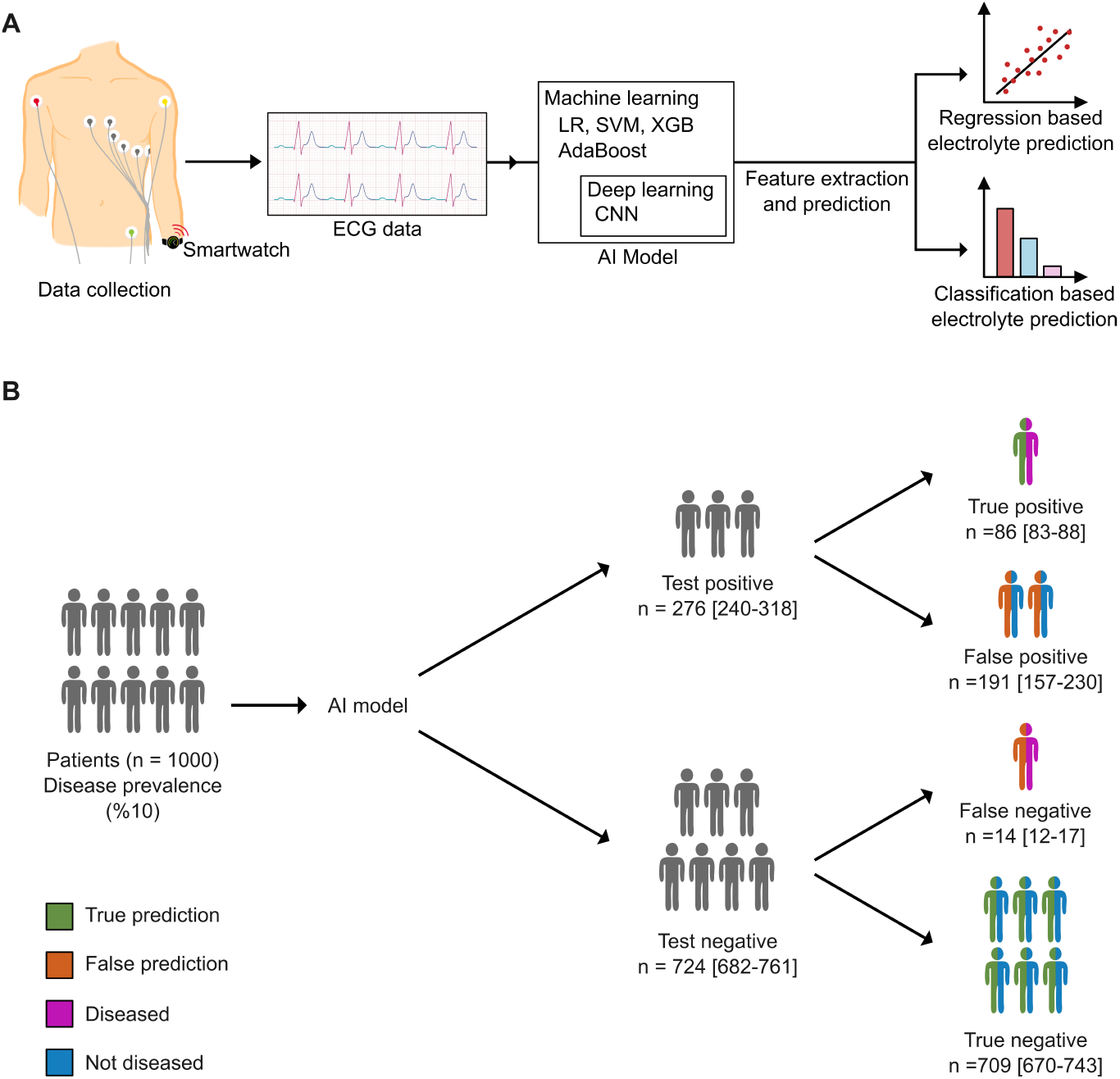
Overall pipeline of studies included in the systematic review. **(A)** Data was collected either through a 12-lead ECG machine in a hospital environment or via a smartwatch. The collected ECGs were processed as either images or signals. Feature extraction was performed using either simple QRS detection algorithms, which involved characterizing each wave with various time and frequency metrics, or automated methods employing deep learning algorithms. The extracted features were then input into traditional machine learning models or advanced deep learning frameworks. Some studies conducted regression-based analyses to predict serum electrolyte levels, while others employed classification approaches to detect abnormalities based on predefined cutoff values. LR: Logistic regression, SVM: Support vector machine, XGB: Extreme gradient boosting, CNN: Convolutional neural networks. **(B)** Hypothetical scenario for deployment of AI model in clinical application for hyperkalemia detection in a cohort of 1000 patients with a 10% disease prevalence.

### Strengths and limitations

This review includes a comprehensive search of databases, encompassing engineering databases and preprints. We evaluated papers in depth and treated each model development as a separate entity to explore the capabilities of AI. We were able to differentiate between internal and external dataset validations, which is a crucial aspect for addressing out-of-domain problems. The main limitation of this study is the number of publications included. Despite an extensive search across 9 different databases, we found only 21 relevant studies for the systematic review, and just 10 of these were included in the meta-analysis. Furthermore, there was a high bias in patient selection in some studies due to unclear data collection methods, non-consecutive sampling, or failure to avoid a case-control design. Additionally, some studies had a low number of training and testing samples, which may have led to overfitting problems that do not accurately reflect the real performance of the models. Model training parameters and epoch-loss graphs for training were not shared in most of the studies.

## Conclusion

Promising AI models stemming from both clinical and engineering fields have demonstrated their potential to enhance ECG-based concentration estimation reliability. The findings in the present study not only affirm the feasibility of ECG-based concentration estimation but also indicate a clear progression toward clinical implementation. However, to integrate such systems into clinical practice further studies should be conducted across diverse clinical settings, hospitals, ethnic groups, countries, and regions to validate the efficacy and generalizability of the models. Furthermore, the lack of comprehensive cost-effectiveness analyses underscores the need for future research to evaluate the economic implications of implementing such smart systems in healthcare settings.

## Data Availability

All data produced in the present work are contained in the manuscript or in the supplementary file

## Notes

### Competing Interest Statement

The authors have declared no competing interest.

### Funding Statement

This study did not receive any funding

### Author Declarations

This systematic review and meta-analysis did not involve the collection of new data from human or animal subjects. All data were retrieved from previously published studies and publicly available datasets. Therefore, no additional ethical approval was required. All data used in the study is included in the manuscript or in supplementary data.

