## Supplementary_file for "AI in ECG-Based Electrolyte Imbalance Prediction: A Systematic Review and Meta Analysis": Supplementary Tables_and_Figures.pdf

**Supplementary Table 1**: Preferred Reporting Items for Systematic Reviews and Meta-analyses of Diagnostic Test Accuracy Studies (PRISMA-DTA) Checklist

| Section/topic | # | PRISMA-DTA Checklist Item | Reported on page # |
|---|---|---|---|
| **TITLE / ABSTRACT** | | | |
| Title | 1 | Identify the report as a systematic review (+/- meta-analysis) of diagnostic test accuracy (DTA) studies. | 1 |
| Abstract | 2 | Abstract: See PRISMA-DTA for abstracts. | 2 |
| **INTRODUCTION** | | | |
| Rationale | 3 | Describe the rationale for the review in the context of what is already known. | 3-4 |
| Clinical role of index test | D1 | State the scientific and clinical background, including the intended use and clinical role of the index test, and if applicable, the rationale for minimally acceptable test accuracy (or minimum difference in accuracy for comparative design). | 3-4 |
| Objectives | 4 | Provide an explicit statement of question(s) being addressed in terms of participants, index test(s), and target condition(s). | 4 |
| **METHODS** | | | |
| Protocol and registration | 5 | Indicate if a review protocol exists, if and where it can be accessed (e.g., Web address), and, if available, provide registration information including registration number. | 5 |
| Eligibility criteria | 6 | Specify study characteristics (participants, setting, index test(s), reference standard(s), target condition(s), and study design) and report characteristics (e.g., years considered, language, publication status) used as criteria for eligibility, giving rationale. | 5 |
| Information sources | 7 | Describe all information sources (e.g., databases with dates of coverage, contact with study authors to identify additional studies) in the search and date last searched. | 5 |
| Search | 8 | Present full search strategies for all electronic databases and other sources searched, including any limits used, such that they could be repeated. | 5 |
| Study selection | 9 | State the process for selecting studies (i.e., screening, eligibility, included in systematic review, and, if applicable, included in the meta-analysis). | 6-7 |

| Section/topic | # | PRISMA-DTA Checklist Item | Reported on page # |
|---|---|---|---|
| Data collection process | 10 | Describe method of data extraction from reports (e.g., piloted forms, independently, in duplicate) and any processes for obtaining and confirming data from investigators. | 6-7 |
| Definitions for data extraction | 11 | Provide definitions used in data extraction and classifications of target condition(s), index test(s), reference standard(s) and other characteristics (e.g. study design, clinical setting). | 6-7 |
| Risk of bias and applicability | 12 | Describe methods used for assessing risk of bias in individual studies and concerns regarding the applicability to the review question. | 6-7 |
| Diagnostic accuracy measures | 13 | State the principal diagnostic accuracy measure(s) reported (e.g. sensitivity, specificity) and state the unit of assessment (e.g. per-patient, per-lesion). | 6-7 |
| Synthesis of results | 14 | Describe methods of handling data, combining results of studies and describing variability between studies. This could include, but is not limited to: a) handling of multiple definitions of target condition. b) handling of multiple thresholds of test positivity, c) handling multiple index test readers, d) handling of indeterminate test results, e) grouping and comparing tests, f) handling of different reference standards | 7 |
| Meta-analysis | D2 | Report the statistical methods used for meta-analyses, if performed. | 7 |
| Additional analyses | 16 | Describe methods of additional analyses (e.g., sensitivity or subgroup analyses, meta-regression), if done, indicating which were pre-specified. | 7 |
| **RESULTS** | | | |
| Study selection | 17 | Provide numbers of studies screened, assessed for eligibility, included in the review (and included in meta-analysis, if applicable) with reasons for exclusions at each stage, ideally with a flow diagram. | 8 |
| Study characteristics | 18 | For each included study provide citations and present key characteristics including: a) participant characteristics (presentation, prior testing), b) clinical setting, c) study design, d) target condition definition, e) index test, f) reference standard, g) sample size, h) funding sources | 8-10 |

| | | | |
|---|---|---|---|
| Risk of bias and applicability | 19 | Present evaluation of risk of bias and concerns regarding applicability for each study. | 10-11 |
| Results of individual studies | 20 | For each analysis in each study (e.g. unique combination of index test, reference standard, and positivity threshold) report 2x2 data (TP, FP, FN, TN) with estimates of diagnostic accuracy and confidence intervals, ideally with a forest or receiver operator characteristic (ROC) plot. | 12-13 |
| Synthesis of results | 21 | Describe test accuracy, including variability; if meta-analysis was done, include results and confidence intervals. | 12-15 |
| Additional analysis | 23 | Give results of additional analyses, if done (e.g., sensitivity or subgroup analyses, meta-regression; analysis of index test: failure rates, proportion of inconclusive results, adverse events). | 12-16 |
| **DISCUSSION** | | | |
| Summary of evidence | 24 | Summarize the main findings including the strength of evidence. | 17-19 |
| Limitations | 25 | Discuss limitations from included studies (e.g. risk of bias and concerns regarding applicability) and from the review process (e.g. incomplete retrieval of identified research). | 19 |
| Conclusions | 26 | Provide a general interpretation of the results in the context of other evidence. Discuss implications for future research and clinical practice (e.g. the intended use and clinical role of the index test). | 19-20 |
| **FUNDING** | | | |
| Funding | 27 | For the systematic review, describe the sources of funding and other support and the role of the funders. | Not applicable |

**Supplementary Table 2:** List of databases screened and corresponding prompts used

| Database | Index and keyword terms |
|---|---|
| PubMed-Medline | ((AI[Title/Abstract] OR artificial intelligence[Title/Abstract] OR machine learning[Title/Abstract] OR deep learning[Title/Abstract] OR neural networks[Title/Abstract] OR computer intelligence[Title/Abstract]) AND (hyperkalemi*[Title/Abstract] OR hypokalemi*[Title/Abstract] OR hyperkalaemi*[Title/Abstract] OR hypokalaemi*[Title/Abstract] OR electrol*[Title/Abstract] OR potass*[Title/Abstract] OR hyperpotass*[Title/Abstract] OR hypopotass*[Title/Abstract] OR calcium[Title/Abstract] OR hypercalcemi*[Title/Abstract] OR hypocalcemi*[Title/Abstract] OR sodium[Title/Abstract] OR hypernatremi*[Title/Abstract] OR hyponatremi*[Title/Abstract])) AND (ecg[Title/Abstract] OR ekg[Title/Abstract] OR electrocardi*[Title/Abstract] OR cardiac electrophysiology[Title/Abstract]) |
| *Web of Science* | (AB=(AI OR artificial intelligence OR machine learning OR deep learning OR neural networks OR computer intelligence) OR TI=(AI OR artificial intelligence OR machine learning OR deep learning OR neural networks OR computer intelligence)) AND (AB=(hyperkalemi* OR hypokalemi* OR hyperkalaemi* OR hypokalaemi* OR electrol* OR potass* OR hyperpotass* OR hypopotass* OR calcium OR hypercalcemi* OR hypocalcemi* OR sodium OR hypernatremi* OR hyponatremi*) OR TI=(hyperkalemi* OR hypokalemi* OR hyperkalaemi* OR hypokalaemi* OR electrol* OR potass* OR hyperpotass* OR hypopotass* OR calcium OR hypercalcemi* OR hypocalcemi* OR sodium OR hypernatremi* OR hyponatremi*)) AND (AB=(ecg OR ekg OR electrocardi* OR cardiac electrophysiology) OR TI=(ecg OR ekg OR electrocardi* OR cardiac electrophysiology)) |
| *Scopus* | TITLE-ABS ( ai OR "artificial intelligence" OR "machine learning" OR "deep learning" OR "neural networks" OR "computer intelligence" ) AND TITLE-ABS ( hyperkalemi* OR hypokalemi* OR hyperkalaemi* OR hypokalaemi* OR electrol* OR potass* OR hyperpotass* OR hypopotass* OR calcium OR hypercalcemi* OR hypocalcemi* OR sodium OR hypernatremi* OR hyponatremi* ) AND TITLE-ABS ( ecg OR ekg OR electrocardi* OR "cardiac electrophysiology" ) |
| *IEEE Explore* | ("All Metadata":AI OR "All Metadata":artificial intelligence OR "All Metadata":machine learning OR "All Metadata":deep learning OR "All Metadata":neural networks OR "All Metadata":computer intelligence) AND ("All Metadata":hyperkalemia OR "All Metadata":hypokalemia OR "All Metadata":hyperkalemic OR "All Metadata":hypokalemic OR "All Metadata":hyperkalaemia OR "All Metadata":hypokalaemia OR "All Metadata":hyperkalaemic OR "All Metadata":hypokalaemic OR "All Metadata":electrol* OR "All Metadata":potass* OR "All Metadata":hyperpotass* OR "All Metadata":hypopotass* OR "All Metadata":calcium OR "All Metadata":hypercalcemi* OR "All Metadata":hypocalcemi* OR "All Metadata":sodium OR "All Metadata":hypernatremi* OR "All Metadata":hyponatremi*) AND ("All |

| | |
|---|---|
| | Metadata":ecg OR "All Metadata":ekg OR "All Metadata":electrocardi* OR "All Metadata":cardiac electrophysiology) |
| *Google Scholar* | (AI OR artificial intelligence OR machine learning OR deep learning OR neural networks OR computer intelligence) AND (ecg OR ekg OR electrocardi* OR cardiac electrophysiology) AND (hyperkalemi* OR hypokalemi* OR hyperkalaemi* OR hypokalaemi* OR electrol* OR potass* OR hyperpotass* OR hypopotass* OR calcium OR hypercalcemi* OR hypocalcemi* OR sodium OR hypernatremi* OR hyponatremi*) |
| *Arxiv&bioarxiv* | order: -announced_date_first; size: 200; include_cross_list: True; terms: AND abstract=AI OR "artificial intelligence" OR "machine learning" OR "deep learning" OR "neural networks" OR "computer intelligence"; AND abstract=ecg OR ekg OR electrocardi* OR "cardiac electrophysiology"; AND abstract=hyperkalemi* OR hypokalemi* OR hyperkalaemi* OR hypokalaemi* OR electrol* OR potass* OR hyperpotass* OR hypopotass* OR calcium OR hypercalcemi* OR hypocalcemi* OR sodium OR hypernatremi* OR hyponatremi* |
| *EBSCO* | bquery=AB+(+AI+OR+artificial+intelligence+OR+machine+learning+OR+deep+learning+OR+neural+networks+OR+computer+intelligence+)+AND+AB+(+hyperkalemi*+OR+hypokalemi*+OR+hyperkalaemi*+OR+hypokalaemi*+OR+electrol*+OR+potass*+OR+hyperpotass*+OR+hypopotass*+OR+calcium+OR+hypercalcemi*+OR+hypocalcemi*+OR+sodium+OR+hypernatremi*+OR+hyponatremi*+)+AND+AB+(+ecg+OR+ekg+OR+electrocardi*+OR+cardiac+electrophysiology+) |
| *The Cochrane Library* | (AI OR artificial intelligence OR machine learning OR deep learning OR neural networks OR computer intelligence) AND (ecg OR ekg OR electrocardi* OR cardiac electrophysiology) AND (hyperkalemi* OR hypokalemi* OR hyperkalaemi* OR hypokalaemi* OR electrol* OR potass* OR hyperpotass* OR hypopotass* OR calcium OR hypercalcemi* OR hypocalcemi* OR sodium OR hypernatremi* OR hyponatremi*) |

**Supplementary Table 3.** Kappa agreement for QUADAS-2

| QUADAS Item | | Kappa |
|---|---|---|
| Overall | | 0.805 |
| Domain 1: Patient Selection | Could the selection of patients have introduced bias? | 0.905 |
| | Concerns regarding applicability: Is there concern that the included patients do not match the review question? | 0.736 |
| Domain 2: Index Test | Could the conduct or interpretation of the index test have introduced bias? | 0.759 |
| | Is there concern that the index test, its conduct, or interpretation differ from the review question? | 0.868 |
| Domain 3: Reference Test | Could the reference standard, its conduct, or its interpretation have introduced bias? | 0.674 |
| | Is there concern that the target condition as defined by the reference standard does not match the review question? | 0.708 |
| Domain 4: Flow and Timing | Could the patient flow have introduced bias? | 0.778 |

**Supplementary Table 4.** Detailed QUADAS-2 assessment of studies included in the systematic review

| Study name | Risk of bias | | | | Applicability | | |
|---|---|---|---|---|---|---|---|
| | Patient selection | Index test | Reference standard | Flow and timing | Patient selection | Index test | Reference standard |
| An J et.al. 2024 | High | Low | Low | High | Low | Low | Low |
| Babur S et. al. 2024 | High | Low | Low | High | High | Low | Low |
| Chiu IM et. al. 2022 | High | Low | Low | High | High | High | Low |
| Chiu IM et. al. 2024 | Low | Low | Low | Low | Low | Low | Low |
| Galloway CD et. al. 2019 | High | Low | Low | High | High | Low | Low |
| Harmon DM et. al. 2024 | Low | Low | Low | Unclear | Low | Low | Low |
| Harmon DM et. al. 2024b | Low | Low | Low | High | Low | Low | Low |
| Khan RU et. al. 2019 | High | Unclear | Unclear | Unclear | High | Unclear | Unclear |
| Kim D et. al. 2023 | High | Low | High | Unclear | High | Low | High |
| Kwon JM et. al. 2021 | Low | Low | Low | Low | Low | Low | Low |
| Lin C et. al. 2022 | Low | Low | Low | Low | Low | Low | Low |
| Lin C et. al. 2022b | Low | Low | Low | Low | Low | Low | Low |
| Lin CS et. al. 2020 | Low | Low | Low | Low | Low | Low | Low |
| Lou YS et. al. 2023 | Low | Low | Low | Low | Low | Low | Low |
| Lou YS et. al. 2023b | Low | Low | High | Unclear | Low | Low | High |
| Urtnasan E et. al. 2022 | High | High | Unclear | Low | High | High | Unclear |
| Von Bachmann P et. al. 2024 | Low | Low | Low | Low | Low | Low | Low |
| Wang CX et. al. 2021 | High | Low | Low | Low | High | Low | Low |
| Wang Y et. al. 2022 | High | Low | Low | High | High | Low | Low |
| Wu MF et. al. 2003 | High | Unclear | Low | Unclear | High | Unclear | Low |
| Xu D et. al. 2023 | High | High | Low | Low | High | High | Low |

**Supplementary Table 5.** Meta-regression analysis of sensitivity measures in hyperkalemia studies.

| Variable | Estimate | SE | Z-value | P-value | CI Lower | CI Upper |
|---|---|---|---|---|---|---|
| intrcpt | -0.052 | 0.905 | -0.058 | 0.954 | -1.826 | 1.721 |
| Leads1 lead | 0.237 | 0.281 | 0.845 | 0.398 | -0.313 | 0.787 |
| Leads2 lead | 0.368 | 0.926 | 0.398 | 0.691 | -1.446 | 2.183 |
| Leads3 lead | 0.011 | 0.352 | 0.031 | 0.975 | -0.680 | 0.702 |
| Leads4 lead | 0.586 | 0.873 | 0.671 | 0.502 | -1.125 | 2.296 |
| Leads6 lead | 0.675 | 0.451 | 1.499 | 0.134 | -0.208 | 1.559 |
| Leads7 lead | -1.638 | 0.836 | -1.960 | 0.050 | -3.277 | 0.000 |
| setexternal test | 0.003 | 0.190 | 0.017 | 0.986 | -0.369 | 0.375 |
| RegionUSA | -0.503 | 0.883 | -0.570 | 0.569 | -2.234 | 1.228 |
| patient_selectionHigh | 0.307 | 0.277 | 1.106 | 0.269 | -0.237 | 0.851 |
| model_typedeep learning | 1.800 | 0.792 | 2.274 | 0.023 | 0.248 | 3.352 |
| sssample size>1000 | -0.183 | 0.449 | -0.407 | 0.684 | -1.063 | 0.698 |

$I^2$ = 94.13%, τ2= 0.2623, τ= 0.5122

Test of moderators: QM(df = 11) = 20.7630, p-val = 0.0359

**Supplementary Table 6.** Meta-regression analysis of specificity measures in hyperkalemia studies.

| Variable | Estimate | SE | Z-value | P-value | CI Lower | CI Upper |
|---|---|---|---|---|---|---|
| intrcpt | 5.005 | 0.854 | 5.864 | 0.000 | 3.332 | 6.678 |
| Leads1 lead | -0.694 | 0.297 | -2.333 | 0.020 | -1.277 | -0.111 |
| Leads2 lead | -3.176 | 0.870 | -3.651 | 0.000 | -4.881 | -1.471 |
| Leads3 lead | -0.275 | 0.378 | -0.728 | 0.466 | -1.016 | 0.466 |

| | | | | | | |
|---|---|---|---|---|---|---|
| Leads4 lead | -3.011 | 0.806 | -3.735 | 0.000 | -4.591 | -1.431 |
| Leads6 lead | -0.875 | 0.458 | -1.912 | 0.056 | -1.771 | 0.022 |
| Leads7 lead | -2.940 | 0.921 | -3.192 | 0.001 | -4.745 | -1.135 |
| setexternal test | -0.328 | 0.201 | -1.629 | 0.103 | -0.723 | 0.067 |
| RegionUSA | 2.395 | 0.819 | 2.923 | 0.003 | 0.789 | 4.000 |
| patient_selectionHigh | -0.163 | 0.293 | -0.558 | 0.577 | -0.737 | 0.410 |
| model_typedeep learning | -1.261 | 0.665 | -1.896 | 0.058 | -2.565 | 0.042 |
| sssample size>1000 | -1.781 | 0.530 | -3.363 | 0.001 | -2.819 | -0.743 |

$I^2$ = 99.93%, τ2= 0.3190 , τ= 0.5648

Test of moderators: QM(df = 11) = 38.6320, p-val < .0001

**Supplementary Table 7.** Meta-regression analysis of diagnostic odds ratio measures in hyperkalemia studies.

| **Variable** | **Estimate** | **SE** | **Z-value** | **P-value** | **CI Lower** | **CI Upper** |
|---|---|---|---|---|---|---|
| intrcpt | 5.234 | 0.859 | 6.091 | 0.000 | 3.550 | 6.919 |
| Leads1 lead | -0.532 | 0.213 | -2.501 | 0.012 | -0.949 | -0.115 |
| Leads2 lead | -2.970 | 0.857 | -3.467 | 0.001 | -4.648 | -1.291 |
| Leads3 lead | -0.244 | 0.259 | -0.942 | 0.346 | -0.753 | 0.264 |
| Leads4 lead | -2.594 | 0.825 | -3.145 | 0.002 | -4.211 | -0.977 |
| Leads6 lead | -0.316 | 0.348 | -0.907 | 0.364 | -0.999 | 0.367 |
| Leads7 lead | -4.708 | 0.900 | -5.229 | 0.000 | -6.473 | -2.943 |
| setexternal test | -0.293 | 0.141 | -2.078 | 0.038 | -0.569 | -0.017 |
| RegionUSA | 2.059 | 0.831 | 2.477 | 0.013 | 0.430 | 3.688 |
| patient_selectionHigh | 0.031 | 0.212 | 0.145 | 0.885 | -0.385 | 0.446 |
| model_typedeep learning | 0.468 | 0.710 | 0.659 | 0.510 | -0.923 | 1.859 |
| sssample size>1000 | -2.083 | 0.486 | -4.289 | 0.000 | -3.035 | -1.131 |

$I^2$ = 88.89%, τ2= 0.1353, τ= 0.3678

Test of moderators: QM(df = 11) = 61.6934, p-val < .0001

**Supplementary Table 8.** Meta-regression analysis of sensitivity measures in hypokalemia studies.

| Variable | Estimate | SE | Z-value | P-value | CI Lower | CI Upper |
|---|---|---|---|---|---|---|
| intrcpt | 1.058 | 0.433 | 2.444 | 0.015 | 0.210 | 1.907 |
| Leads1 lead | 0.218 | 0.267 | 0.816 | 0.414 | -0.305 | 0.741 |
| Leads3 lead | 0.144 | 0.348 | 0.415 | 0.678 | -0.537 | 0.826 |
| Leads6 lead | 0.755 | 0.459 | 1.645 | 0.100 | -0.145 | 1.654 |
| setexternal test | -0.365 | 0.226 | -1.618 | 0.106 | -0.807 | 0.077 |
| patient_selectionHigh | -0.119 | 0.260 | -0.458 | 0.647 | -0.630 | 0.391 |
| sssample size>1000 | 0.637 | 0.462 | 1.378 | 0.168 | -0.269 | 1.544 |

$I^2$ = 99.10%, τ2= 0.3099, τ= 0.5567

Test of moderators: QM(df = 6) = 8.4424, p-val = 0.2074

**Supplementary Table 9.** Meta-regression analysis of specificity measures in hypokalemia studies.

| Variable | Estimate | SE | Z-value | P-value | CI Lower | CI Upper |
|---|---|---|---|---|---|---|
| intrcpt | 1.457 | 0.407 | 3.579 | 0.000 | 0.659 | 2.255 |
| Leads1 lead | -0.610 | 0.240 | -2.541 | 0.011 | -1.080 | -0.139 |
| Leads3 lead | 0.079 | 0.314 | 0.253 | 0.800 | -0.535 | 0.694 |
| Leads6 lead | -0.576 | 0.409 | -1.410 | 0.158 | -1.378 | 0.225 |
| setexternal test | -0.013 | 0.203 | -0.062 | 0.951 | -0.410 | 0.385 |
| patient_selectionHigh | 0.598 | 0.235 | 2.545 | 0.011 | 0.137 | 1.058 |

| | | | | | | |
|---|---|---|---|---|---|---|
| sssample size>1000 | -0.702 | 0.430 | -1.634 | 0.102 | -1.544 | 0.140 |

$I^2 = 99.93\%$, $\tau2= 0.2544$, $\tau= 0.5044$

Test of moderators: QM(df = 6) = 24.2571, p-val = 0.0005

**Supplementary Table 10.** Meta-regression analysis of diagnostic odds ratio measures in hypokalemia studies.

| Variable | Estimate | SE | Z-value | P-value | CI Lower | CI Upper |
|---|---|---|---|---|---|---|
| intrcpt | 2.554 | 0.611 | 4.181 | 0.000 | 1.357 | 3.751 |
| Leads1 lead | -0.388 | 0.368 | -1.055 | 0.291 | -1.109 | 0.333 |
| Leads3 lead | 0.230 | 0.480 | 0.480 | 0.631 | -0.710 | 1.170 |
| Leads6 lead | 0.162 | 0.629 | 0.258 | 0.797 | -1.071 | 1.396 |
| setexternal test | -0.377 | 0.311 | -1.214 | 0.225 | -0.986 | 0.232 |
| patient_selectionHigh | 0.462 | 0.359 | 1.286 | 0.198 | -0.242 | 1.166 |
| sssample size>1000 | -0.093 | 0.648 | -0.143 | 0.886 | -1.363 | 1.177 |

$I^2 = 99.46\%$, $\tau2= 0.5923$, $\tau= 0.7696$

Test of moderators: QM(df = 6) = 5.9808, p-val = 0.4253

**Supplementary Table 11.** Baujat graphical method x and y coordinates for studies contributing to the meta-analysis of hyperkalemia detection. Contribution to heterogeneity and influence on overall results decrease as the coordinates approach the origin.

| Study Sub-Name | Contribution_to_Heterogeneity | Influence_on_Overall_Results |
|---|---|---|
| An JN et. al. 2024 (S1) | 3.197695564 | 0.045890074 |
| An JN et. al. 2024 (S2) | 15.71562585 | 0.378417988 |
| An JN et. al. 2024 (S3) | 0.613979965 | 0.009018415 |
| An JN et. al. 2024 (S4) | 1.844731941 | 0.050634099 |

| | | |
|---|---|---|
| An JN et. al. 2024 (S5) | 3.295672871 | 0.058074686 |
| An JN et. al. 2024 (S6) | 1.599401365 | 0.0481869 |
| An JN et. al. 2024 (S7) | 6.225697878 | 0.308767718 |
| An JN et. al. 2024 (S8) | 46.12128708 | 3.950729949 |
| An JN et. al. 2024 (S9) | 1.500837892 | 0.07995374 |
| An JN et. al. 2024 (S10) | 9.361633693 | 0.913861456 |
| An JN et. al. 2024 (S11) | 12.40562179 | 0.734574025 |
| An JN et. al. 2024 (S12) | 7.379726723 | 0.761418209 |
| Galloway CD et. al. 2019 (S1) | 14.58181459 | 1.19124828 |
| Galloway CD et. al. 2019 (S2) | 14.53967048 | 0.672758688 |
| Galloway CD et. al. 2019 (S3) | 0.015377585 | 0.001146093 |
| Galloway CD et. al. 2019 (S4) | 2.030040096 | 0.093857201 |
| Galloway CD et. al. 2019 (S5) | 20.18539773 | 0.367594371 |
| Galloway CD et. al. 2019 (S6) | 5.528616844 | 0.04823101 |
| Galloway CD et. al. 2019 (S7) | 2.985873305 | 0.044874258 |
| Galloway CD et. al. 2019 (S8) | 0.844944605 | 0.007099863 |
| Galloway CD et. al. 2019 (S9) | 22.00069684 | 0.388918981 |
| Galloway CD et. al. 2019 (S10) | 21.51800903 | 0.237665852 |
| Galloway CD et. al. 2019 (S11) | 3.960454718 | 0.057787204 |
| Galloway CD et. al. 2019 (S12) | 0.262992239 | 0.001947085 |
| Kwon JM et. al. 2021 (S1) | 15.82028474 | 0.136403203 |
| Kwon JM et. al. 2021 (S2) | 13.65577347 | 0.105185532 |
| Kwon JM et. al. 2021 (S3) | 16.53431988 | 0.162056126 |
| Kwon JM et. al. 2021 (S4) | 5.360291305 | 0.037151764 |
| Kwon JM et. al. 2021 (S5) | 9.936743177 | 0.071912041 |
| Kwon JM et. al. 2021 (S6) | 30.23026991 | 0.20952825 |
| Lin C et. al. 2022 (S1) | 68.35713703 | 2.157862774 |
| Lin C et. al. 2022 (S4) | 10.52179537 | 0.035728512 |
| Lin CS et. al. 2020 (S1) | 6.812272738 | 0.0089447 |
| Lin CS et. al. 2020 (S3) | 0.937524557 | 0.029393818 |
| Urtnasan E et. al. 2022 | 35.72965294 | 0.044242466 |
| Wu MF et. al. 2003 | 10.06849756 | 0.009064791 |

| | | |
|---|---|---|
| Xu D et. al. 2023 (S2) | 0.004753657 | 6.15033E-06 |
| Xu D et. al. 2023 (S6) | 0.552018246 | 0.001443459 |
| Xu D et. al. 2023 (S10) | 0.636091216 | 0.00169731 |
| Xu D et. al. 2023 (S14) | 2.575915326 | 0.006956638 |
| Xu D et. al. 2023 (S18) | 1.816122757 | 0.004647717 |

**Supplementary Table 12.** Baujat graphical method x and y coordinates for studies contributing to the meta-analysis of hypokalemia detection. Contribution to heterogeneity and influence on overall results decrease as the coordinates approach the origin.

| **Study Sub-Name** | **Contribution_to_Heterogeneity** | **Influence_on_Overall_Results** |
|---|---|---|
| An JN et. al. 2024 (S13) | 126.3413088 | 1.589904145 |
| An JN et. al. 2024 (S14) | 119.6591666 | 1.433136698 |
| An JN et. al. 2024 (S15) | 37.48517311 | 0.548547784 |
| An JN et. al. 2024 (S16) | 31.07105204 | 0.606398781 |
| An JN et. al. 2024 (S17) | 21.7050635 | 0.297113283 |
| An JN et. al. 2024 (S18) | 14.31004019 | 0.268492929 |
| An JN et. al. 2024 (S19) | 338.2040113 | 39.89473773 |
| An JN et. al. 2024 (S20) | 334.67933 | 37.60821108 |
| An JN et. al. 2024 (S21) | 26.47251816 | 3.245072777 |
| An JN et. al. 2024 (S22) | 28.64007284 | 4.141652049 |
| An JN et. al. 2024 (S23) | 0.031991387 | 0.00383572 |
| An JN et. al. 2024 (S24) | 145.299065 | 17.15705833 |
| Kwon JM et. al. 2021 (S7) | 7.486842719 | 0.05432225 |
| Kwon JM et. al. 2021 (S8) | 0.698630374 | 0.00492781 |
| Kwon JM et. al. 2021 (S9) | 2.226869303 | 0.01102442 |
| Kwon JM et. al. 2021 (S10) | 6.719422798 | 0.064139102 |
| Kwon JM et. al. 2021 (S11) | 25.08318809 | 0.201776546 |
| Kwon JM et. al. 2021 (S12) | 74.19708445 | 0.759196608 |
| Lin C et. al. 2022 (S9) | 1139.635436 | 135.5144425 |
| Lin C et. al. 2022 (S12) | 260.806185 | 4.935677867 |
| Lin CS et. al. 2020 (S5) | 3.928773736 | 0.003028198 |
| Lin CS et. al. 2020 (S7) | 660.6049321 | 34.28786084 |

| | | |
|---|---|---|
| Wang CX et. al. 2021 (S1) | 23.6249538 | 0.190942012 |
| Wang CX et. al. 2021 (S2) | 297.8878769 | 2.902450793 |
| Wang CX et. al. 2021 (S3) | 70.05413261 | 0.468234109 |
| Wang CX et. al. 2021 (S4) | 216.6481449 | 1.544671266 |
| Wang Y et. al. 2022 | 3.32559509 | 0.008218127 |

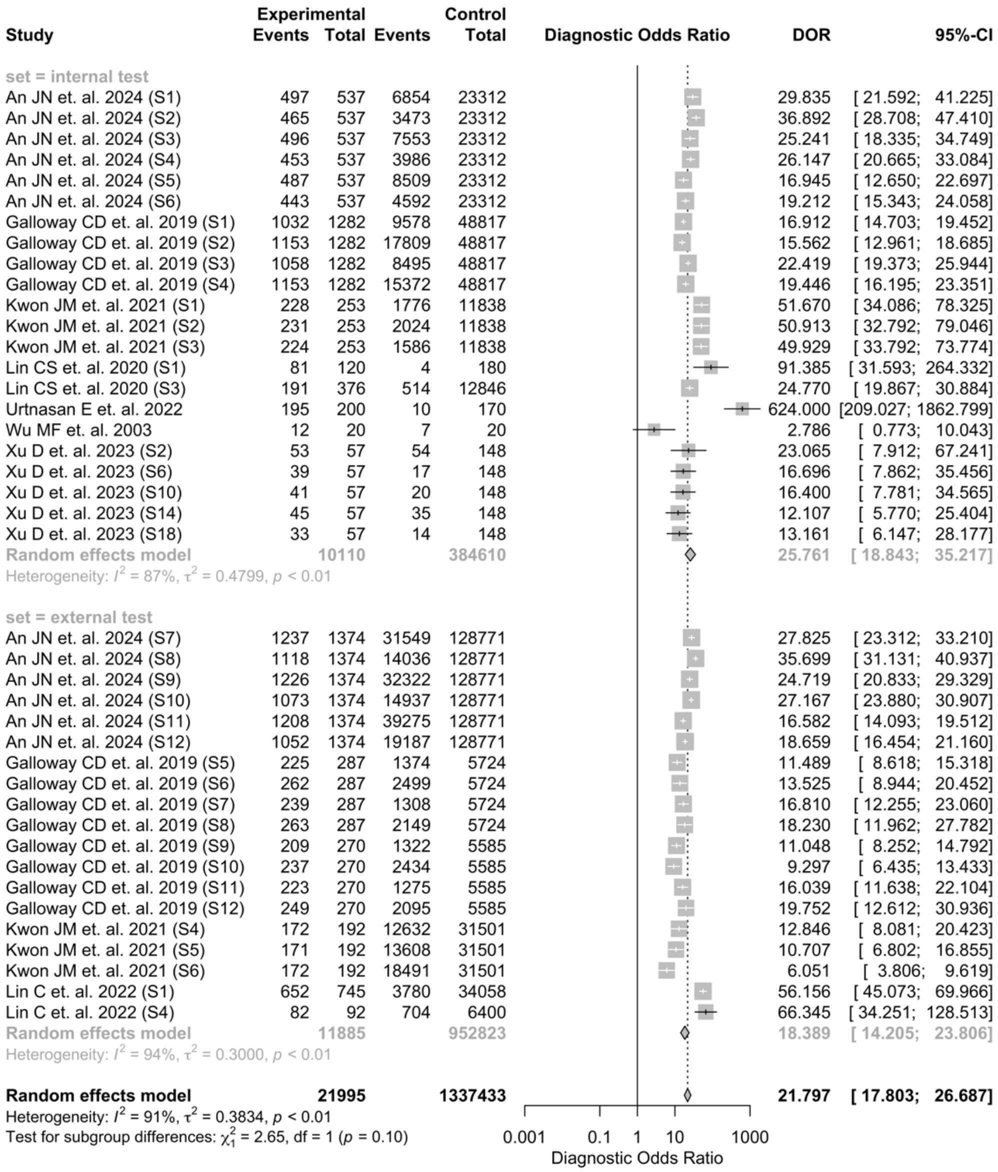


**Supplementary Figure 1.** Pooled diagnostic odds ratio of hyperkalemia detection

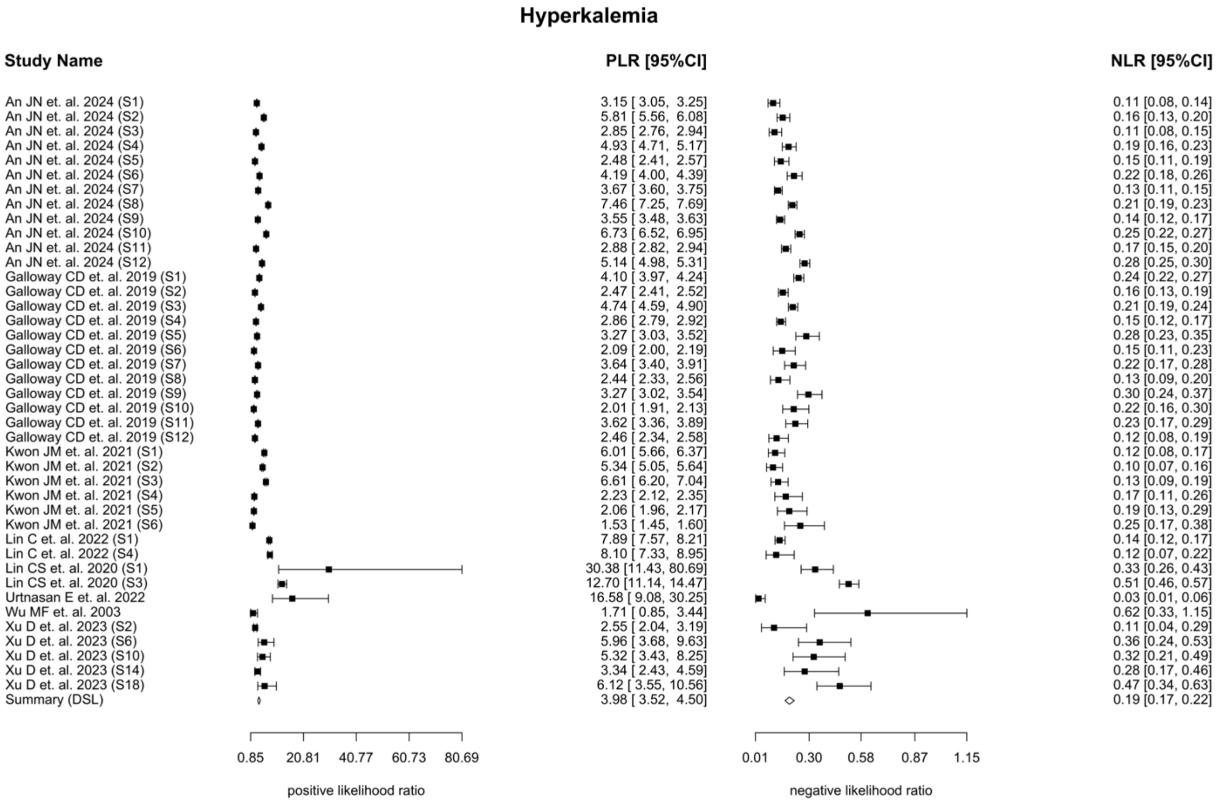


**Supplementary Figure 2.** Pooled positive and negative likelihood ratio for hyperkalemia detection

| Study | Experimental Events | Experimental Total | Control Events | Control Total | DOR | 95%-CI |
|---|---|---|---|---|---|---|
| **set = internal test** | | | | | | |
| An JN et. al. 2024 (S13) | 1489 | 1633 | 4665 | 22216 | 38.903 | [32.688; 46.300] |
| An JN et. al. 2024 (S14) | 1497 | 1633 | 4910 | 22216 | 38.797 | [32.459; 46.373] |
| An JN et. al. 2024 (S15) | 1463 | 1633 | 5909 | 22216 | 23.750 | [20.206; 27.914] |
| An JN et. al. 2024 (S16) | 1391 | 1633 | 4710 | 22216 | 21.364 | [18.568; 24.580] |
| An JN et. al. 2024 (S17) | 1476 | 1633 | 6798 | 22216 | 21.322 | [18.043; 25.197] |
| An JN et. al. 2024 (S18) | 1404 | 1633 | 5443 | 22216 | 18.893 | [16.376; 21.797] |
| Kwon JM et. al. 2021 (S7) | 705 | 790 | 3345 | 11301 | 19.727 | [15.695; 24.795] |
| Kwon JM et. al. 2021 (S8) | 708 | 790 | 3989 | 11301 | 15.827 | [12.551; 19.957] |
| Kwon JM et. al. 2021 (S9) | 735 | 790 | 6046 | 11301 | 11.615 | [ 8.810; 15.314] |
| Lin CS et. al. 2020 (S5) | 81 | 120 | 12 | 180 | 29.077 | [14.450; 58.508] |
| Lin CS et. al. 2020 (S7) | 1519 | 2996 | 1882 | 10226 | 4.560 | [ 4.178; 4.976] |
| Wang CX et. al. 2021 (S1) | 616 | 863 | 198 | 863 | 8.376 | [ 6.744; 10.403] |
| Wang CX et. al. 2021 (S2) | 604 | 863 | 415 | 863 | 2.517 | [ 2.066; 3.067] |
| Wang Y et. al. 2022 | 214 | 276 | 71 | 276 | 9.966 | [ 6.742; 14.732] |
| **Random effects model** | | **17286** | | **179607** | **14.981** | **[ 9.896; 22.677]** |
| Heterogeneity: $I^2 = 99\%$, $\tau^2 = 0.6078$, $p < 0.01$ | | | | | | |
| **set = external test** | | | | | | |
| An JN et. al. 2024 (S19) | 5580 | 7054 | 16125 | 123091 | 25.112 | [23.656; 26.658] |
| An JN et. al. 2024 (S20) | 5678 | 7054 | 17233 | 123091 | 25.348 | [23.846; 26.944] |
| An JN et. al. 2024 (S21) | 5544 | 7054 | 22156 | 123091 | 16.726 | [15.772; 17.738] |
| An JN et. al. 2024 (S22) | 5093 | 7054 | 16617 | 123091 | 16.641 | [15.757; 17.575] |
| An JN et. al. 2024 (S23) | 5601 | 7054 | 25972 | 123091 | 14.414 | [13.584; 15.295] |
| An JN et. al. 2024 (S24) | 5601 | 7054 | 19325 | 123091 | 20.698 | [19.499; 21.972] |
| Kwon JM et. al. 2021 (S10) | 943 | 1052 | 13482 | 30641 | 11.011 | [ 9.019; 13.443] |
| Kwon JM et. al. 2021 (S11) | 962 | 1052 | 17312 | 30641 | 8.230 | [ 6.623; 10.226] |
| Kwon JM et. al. 2021 (S12) | 934 | 1052 | 17251 | 30641 | 6.144 | [ 5.066; 7.450] |
| Lin C et. al. 2022 (S9) | 5624 | 7313 | 10804 | 27490 | 5.143 | [ 4.845; 5.458] |
| Lin C et. al. 2022 (S12) | 849 | 1151 | 2072 | 5341 | 4.435 | [ 3.847; 5.114] |
| Wang CX et. al. 2021 (S3) | 447 | 639 | 198 | 639 | 5.185 | [ 4.087; 6.580] |
| Wang CX et. al. 2021 (S4) | 453 | 639 | 313 | 639 | 2.537 | [ 2.014; 3.195] |
| **Random effects model** | | **55222** | | **864578** | **9.954** | **[ 6.655; 14.889]** |
| Heterogeneity: $I^2 = 100\%$, $\tau^2 = 0.5431$, $p = 0$ | | | | | | |
| **Random effects model** | | **72508** | | **1044185** | **12.274** | **[ 9.148; 16.468]** |
| Heterogeneity: $I^2 = 99\%$, $\tau^2 = 0.5950$, $p = 0$ | | | | | | |
| Test for subgroup differences: $\chi^2_1 = 1.92$, df = 1 ($p = 0.17$) | | | | | | |

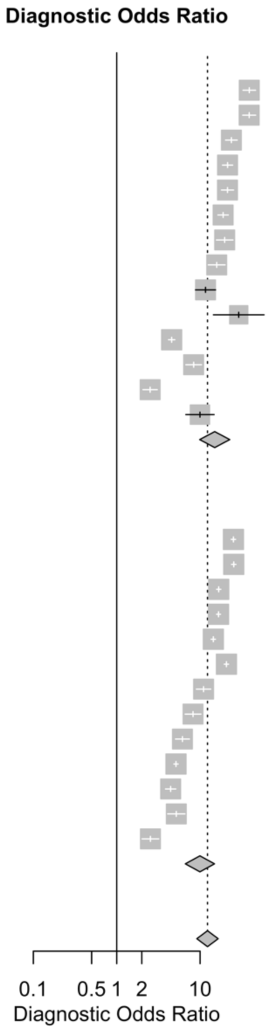


**Supplementary Figure 3.** Pooled diagnostic odds ratio of hypokalemia detection

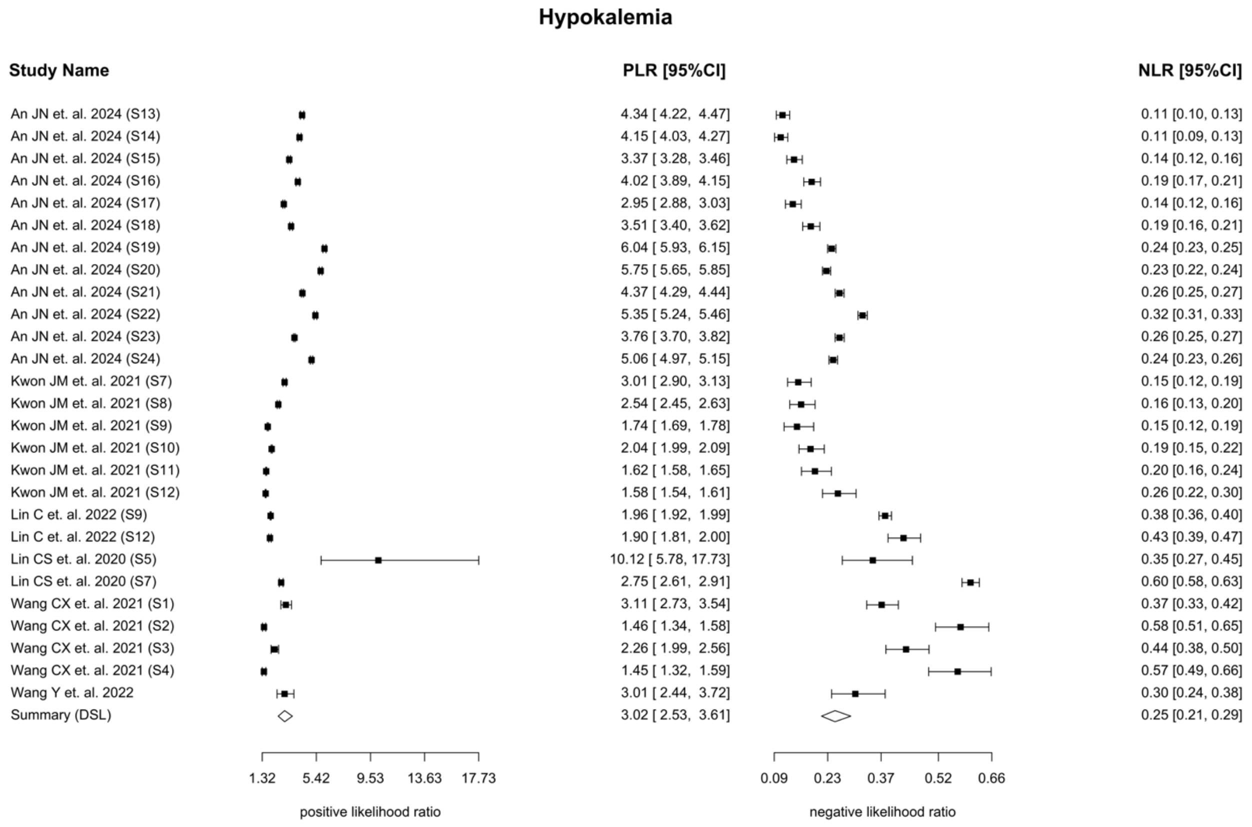


**Supplementary Figure 4.** Pooled positive and negative likelihood ratio for hypokalemia detection

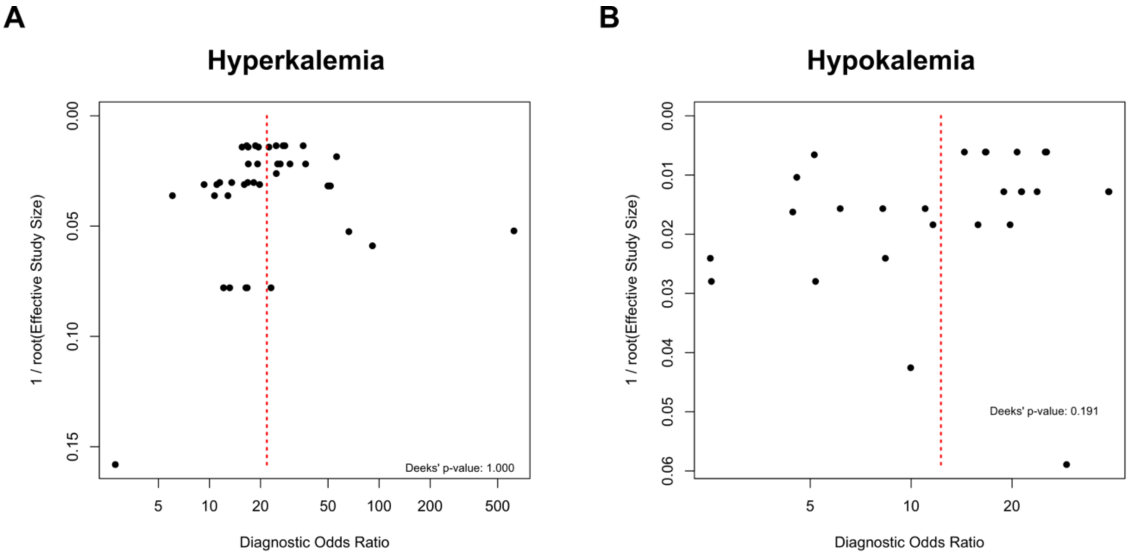


**Supplementary Figure 5.** Duke's bias plot for (A) hyperkalemia, (B) hypokalemia
